# Controlled human influenza infection reveals heterogeneous expulsion of infectious virus into air

**DOI:** 10.1101/2025.11.03.25339190

**Authors:** Nahara Vargas-Maldonado, Nishit Shetty, Lucas M. Ferreri, Matthew D. Pauly, Kayle Patatanian, Shamika Danzy, Meredith J. Shephard, David VanInsberghe, Michelle N. Vu, A.J. Campbell, Kayla Brizuela, Vedhika Raghunathan, Jin Pan, Aaron J. Prussin, Anna Sims, Hollie Macenczak, Jessica Traenkner, Ralph Tanios, Christelle Radi, Veronica Smith, Dalia Gulick, Katia Koelle, Andrew Catchpole, Alex Mann, Colleen S. Kraft, Nadine G. Rouphael, Linsey C. Marr, Anice C. Lowen, Seema S. Lakdawala

## Abstract

Influenza virus is transmitted via respiratory expulsions, but detection of infectious virus in such expulsions has been challenging. Here, we describe quantification and genotyping of infectious virus in respiratory particles using a Modular Influenza Sampling Tunnel (MIST). The particles deposit on cell monolayers, enabling culture, quantification, and sequencing of viruses. Concomitantly, water-sensitive paper and fine particle samplers yield respiratory particle counts over a broad size range. Using the MIST, we captured infectious virus from humans experimentally infected with influenza virus on multiple days post-inoculation. The recovered respiratory particles varied in quantity over three orders of magnitude and contained viral genetic variation that was also detected in samples from infected individuals. Expulsion of infectious virus was associated with infectious viral load in saliva and nasopharyngeal swabs and with clinical symptoms. These data reveal the maintenance of viral diversity in expelled aerosols and suggest that heterogeneity among individuals in the magnitude of infectious expulsions may impact forward transmission potential.

## Introduction

Influenza viruses replicate in the epithelium of the respiratory tract and are shed into respiratory secretions lining the mucosa of these tissues. These respiratory secretions thereby become the vehicle that mediates the spread of influenza viruses between hosts. The extent to which this transfer occurs via airborne infectious respiratory particles has received much attention, since interrupting transmission through the air or via contaminated surfaces requires distinct interventions^1^.

For decades, technical challenges in detecting airborne viruses have hindered understanding of how infectious respiratory particles contribute to transmission. Influenza virus RNA is readily detected in the air in hospitals^2–7^ and other settings^8–10^ and some studies have detected infectious virus in the air^11–15^. One such study found that 70% (37/53) of participants who were positive for influenza A virus (IAV) infection released particles containing culturable virus^14^. While an important advance, this work did not quantify the amount of infectious respiratory particles released by an infected individual and could not sample particles larger than ∼10 μm. Another study quantified infectious virus within fine particles (<5 μm) and reported detection in 39% (55/142) of individuals with confirmed IAV infection^15^. Despite a foundation of evidence indicating that influenza viruses are released into the air within respiratory particles, gaps remain that are critical for defining modes of transmission and guiding the use of non-pharmaceutical interventions. Owing to technical difficulties, the viral content of respiratory particles >10 μm in diameter is understudied, despite these comprising a large fraction of respiratory particles by volume^16^. More generally, the relationship between aerosol particle size and infectious viral content remains unknown and is critical to inform intervention strategies.

The transfer of viral variants from one host to another during transmission has important implications for viral evolution: to contribute to viral evolution at a community scale, variants must be transmitted onward. Natural within-host IAV infections typically have low genetic diversity^17,18^. Inferences based on epidemiologically linked infections have furthermore indicated that transmission bottleneck sizes are very small^17,19^. Thus, few independent viral lineages establish in a new host following transmission. It has not been resolved in human infections, however, whether these small bottlenecks result from expulsion being limited to genetically more homogeneous viral populations, or whether these bottlenecks occur at a later stage of the transmission event. The stage at which the bottleneck acts is important to defining the potential for selection to operate during transmission. Evidence from a guinea pig model revealed a major constriction of diversity during the establishment of infection^20^, an observation that is consistent with the selection of beneficial variants during transmission in ferrets^21^. Thus, data from animal models suggest that bottlenecking may occur late in the transmission process, leaving a window of opportunity for selective forces to act on variants with differential fitness.

Here, we quantified total and infectious respiratory particles produced by individuals experimentally inoculated with IAV. We examined how the magnitude and genetic composition of expelled virus populations related to those observed within hosts. Our data reveal the production of infectious respiratory particles carrying substantial viral diversity during several days after inoculation, with the magnitude of these infectious expulsions varying widely by individual.

## Results

Between July 2023 and March 2025, 24 adult volunteers participated in a controlled human challenge study based on protocol-defined eligibility criteria, including having a hemagglutination inhibition titer ≤1:40 against the challenge strain (see Materials and Methods) (Supplementary Fig. 1). Ten eligible participants were inoculated intranasally with 5.5 log_10_ median tissue culture infective dose (TCID_50_) of influenza A/Perth/16/2009 (H3N2) virus using a mucosal atomization device^22–24^. Fourteen participants were inoculated by inhalation of aerosols containing between 2×10^3^ and 3×10^4^ plaque forming units (PFU)^25^.

Eight of the ten intranasally inoculated participants and six of the fourteen participants inoculated by aerosol inhalation exhibited PCR-confirmed infection on >1 day and were further analyzed for expulsion of infectious respiratory particles (Supplementary Fig. 2). To monitor respiratory and systemic symptoms, participants answered the FLU-PRO^©^ questionnaire daily (Supplementary Fig. 3)^26^. Symptoms with the potential to influence expulsion of infectious virus, such as nasal congestion, sneezing, coughing, fatigue, and throat discomfort, were experienced with varied duration and intensity among the participants (Supplementary Fig. 4). No severe symptoms and no complications were noted.

### Modular Inffuenza Sampling Tunnel (MIST) enables characterization of respiratory particles

To characterize expelled infectious respiratory particles, we developed a sampling platform that enables concurrent air sampling and viral culturing. The MIST, based on a tool originally developed for use with a ferret model^27^, was designed to sample respiratory expulsions from humans over a wide range of particle sizes (Fig. 1, Supplementary Fig. 5). Large respiratory particles settle onto cell culture plates positioned along the length of the MIST. The sizes of respiratory particles are assessed using water-sensitive strips located in between the plates. At the far end of the MIST, two instruments sample aerosol particles that are too small to settle, while also generating a steady flow rate from the entrance to the far end of the tunnel. One of these instruments is an AeroTrak, which counts particles that are 0.3 to 25 µm in diameter. In the sampling conducted on intranasally inoculated participants, the second instrument is a BioSpot VIVAS, which captures particles through condensation followed by deposition into a liquid substrate. In the sampling conducted on participants inoculated via aerosol inhalation, the second instrument is a pump which pulled air through an open-face polytetrafluoroethylene (PTFE) membrane. Samples from both the BioSpot VIVAS and the PTFE membrane are processed for detection of infectious virus. Thus, the MIST allows quantification of both total particles and infectious respiratory particles produced by human participants throughout the course of infection.

**Figure 1.**
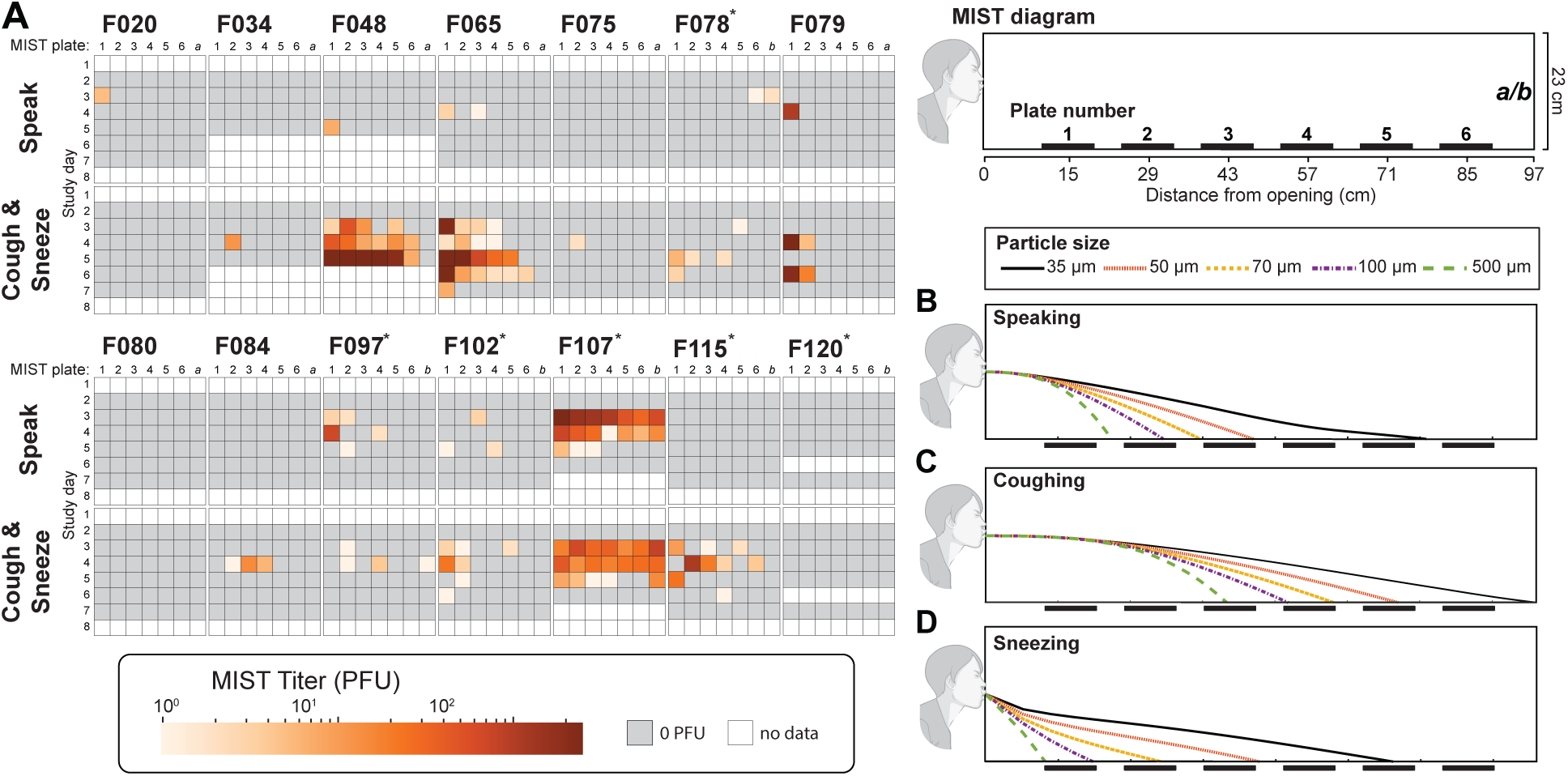
The Modular Influenza Sampling Tunnel (MIST) allows characterization of respiratory particles produced by humans during expiratory activities. The MIST is a cuboidal, static-resistant acrylic tunnel with dimensions of 97 cm (length) x 23 cm (width) x 23 cm (height). (A) Infectious virus detection across cell culture plates 1–6 and aerosol samplers (*a/b*) in the MIST. “a” and “b” represent the BioSpot VIVAS or the Gilair12 pump with open-face PTFE filter, respectively. Participants are denoted with a unique identifier (e.g. F020). Identifiers with an asterisk highlight individuals inoculated via aerosol inhalation. Each frame shows plaque forming units (PFU) detected in MIST samples during speaking or coughing and sneezing, with columns showing plate position and rows showing study day. Squares are colored by the total number of PFU detected. Grey indicates an absence of PFU in the MIST. White that indicates MIST sampling was not done on that day (nd).Predicted respiratory particle deposition within the MIST during (B) speaking, (C) coughing, and (D) sneezing is shown for five particle sizes.

The sampling capabilities of the MIST were validated by generating synthetic aerosols of viral preparations with known infectivity. Three aerosol generating devices were used and, for each, the recovery of infectious virus, viral genomes, and total particles in the MIST was evaluated (Supplementary Fig. 6). Importantly, results indicate reproducibility across three replicate sampling sessions. Infectious virus was detected on the majority of plates but not in the aerosol sampler when virus was delivered in large particles (30–100 µm). Infectious virus was detected only in the aerosol sampler when delivered in particles of 10 µm diameter and was detected only in the source-proximal plates of the MIST when using a jet nebulizer to produce particles with <2 µm diameter. Since particles of <2 µm diameter would not be expected to deposit on these proximal plates, we infer that this positivity results from the low-level production of large droplets during the nebulization process. While positivity across conditions was seen more broadly when testing for viral genomes by RT qPCR, these results indicate that the MIST plates are well-suited to the detection of infectious respiratory particles in the range of 30–100 µm. We note, however, that the trajectories of particles expelled by the synthetic aerosolization devices employed is likely to differ from those produced in natural expulsion events such as coughing and sneezing.

The efficiency of collection of infectious respiratory particles using the BioSpot VIVAS and a PTFE membrane was further examined separately from the MIST tunnel. Each was used to collect infectious aerosols delivered into a 4 L chamber by the jet nebulizer or the flow-focusing monodisperse aerosol generator (FMAG), which produce particles mostly <2 µm and ∼10 µm, respectively. Results indicate that the BioSpot Vivas recovered ∼5% and ∼0.03% of infectious units aerosolized from the nebulizer and the FMAG, respectively, while the PTFE filter allowed recovery of ∼1% of infectious units from both aerosol sources (Supplementary Fig. 7).

### Expulsion of infectious virus varies with individual and is highest during coughing and sneezing

The MIST was used to quantify the amount of infectious influenza virus expelled from the participants during different expiratory activities. First, sampling was performed over a 10-minute period during which participants performed vocal activities including speaking, singing, and spelling. Next, the MIST was reset with fresh plates, and participants mimicked ten coughs and three sneezes. The amount of infectious respiratory particles produced varied widely among individuals (Fig. 1A). Infectious virus in MIST samples was detected as early as study day 3 (2 days post-challenge) and up to study day 7 (6 days post-challenge). For all individuals except F020 and F107, coughing and sneezing led to detection of more infectious virus in MIST samples than did speaking. Notably, for participants F048, F065, and F107, coughing and sneezing led to the deposition of infectious respiratory particles up to the last plate in the MIST, a distance of 85 cm. Most infectious virus was detected closer to the source, however, in line with the observed distribution of total particles detected on water sensitive strips (Supplementary Fig. 8A). Detection of infectious virus in the fine aerosol samples collected at the distal end of the MIST was less common than in the cell plates, with only three participants showing positivity – F078, F097 and F107 (Figure 1A). In addition, influenza virus genetic material (2.6×10^3^ genome copies) was captured by the BioSpot Vivas in the coughing and sneezing sample collected on study day 5 from F048.

### Particle size and number varied among individuals

To model the fate of particles of varying sizes generated during different respiratory activities within the MIST, we employed a transport model that incorporates aerosol dynamics, including particle size, evaporation, and heat transfer (Fig. 1B-D). Based on previously defined characteristics of expiratory events^28^, the modeled expulsions during coughing and speaking included only a horizontal initial velocity, while those during sneezing included an additional downward component. The model predicted that particles would travel farther in the MIST during coughing than during speaking. However, due to their vertical velocity, particles expelled during sneezing were predicted to deposit closer to the source compared to those expelled during speaking. Empirical data on particle size and settling distance was collected during participant sampling sessions from water-sensitive strips placed along the MIST. As predicted by the model, larger diameter particles preferentially deposited closer to the source (Supplementary Fig. 8A). A greater number of particles were detected further from the source during coughing and sneezing compared to speaking (Supplementary Fig. 8A and B), as expected based on the model predictions for coughing. Although the number of particles produced during different expiratory events cannot be compared directly due to different durations of sampling, particle generation within a given expiration type varied among individuals and by day (Supplementary Fig. 8B, C). Production of fine particles also showed marked variation among participants, and a higher concentration of particles was detected during coughing and sneezing than during speaking (paired t-test, p<0.01, n = 98) (Supplementary Fig. 8D) with particles less than 1 μm making up the majority of the particles that reach the distal end of the MIST (Supplementary Fig. 8E).

### The total volume of particles was associated with infectious content

Due to their larger volume, coarse respiratory particles have the potential to carry greater amounts of viral material compared to fine particles. To test whether larger volume corresponds to higher infectivity, we used a linear mixed effects model to evaluate the relationship between the volume of expelled particles and viral titer detected in the MIST, using the total droplet volume on each water-sensitive strip and the PFU detected on the adjacent plate. Analysis of all plates from days and activities when PFU were detected from participants showed significant associations between larger droplet volumes on the water sensitive strips and higher numbers of PFU detected on the plates for both the speaking and the coughing and sneezing activities, suggesting expulsion volume and infectious load are related (Supplementary Fig. 9). Despite this association, for some samples in which infectious virus was detected, the volume of droplets on adjacent strips fell below the detection threshold, indicating that the water-sensitive strips may be ineffective for detecting smaller respiratory particles and that the size of the particles carrying infectious virus may vary by individual. For instance, high levels of PFU were detected from F107 during both the speaking and the coughing and sneezing activities despite low or undetectable droplet volumes.

### Viral load kinetics and spatial distribution differed among inoculated individuals

To assess viral loads and the distribution of viral populations across distinct locations within the nasal and oral cavities, we collected saliva and swabs from the nasopharyngeal site, anterior nasal site, and the oral cavity. Heterogeneity among individuals was apparent in both overall viral loads and the anatomical distribution of the viral population, even among participants inoculated using the same method. Infectious virus was detected in thirteen of the 14 PCR positive individuals tested using the MIST. Each of these 13 participants had infectious virus in nasopharyngeal swab samples, with peak loads differing among individuals by three orders of magnitude. Eight participants had detectable infectious virus in their saliva (Fig. 2). Saliva titers in these participants ranged from 1×10^3^ PFU/mL to >1×10^5^ PFU/mL. The duration of infectious virus detection at any site ranged from 2 days to 7 days, with peak titers typically observed on study days 4 or 5 (days 3 or 4 post-challenge). Expulsion of infectious respiratory particles was observed in 12 of the 13 participants who had infectious virus in their nasopharyngeal swab, saliva samples, or both (Fig. 2 and Supplemental Fig. 2).

**Figure 2.**
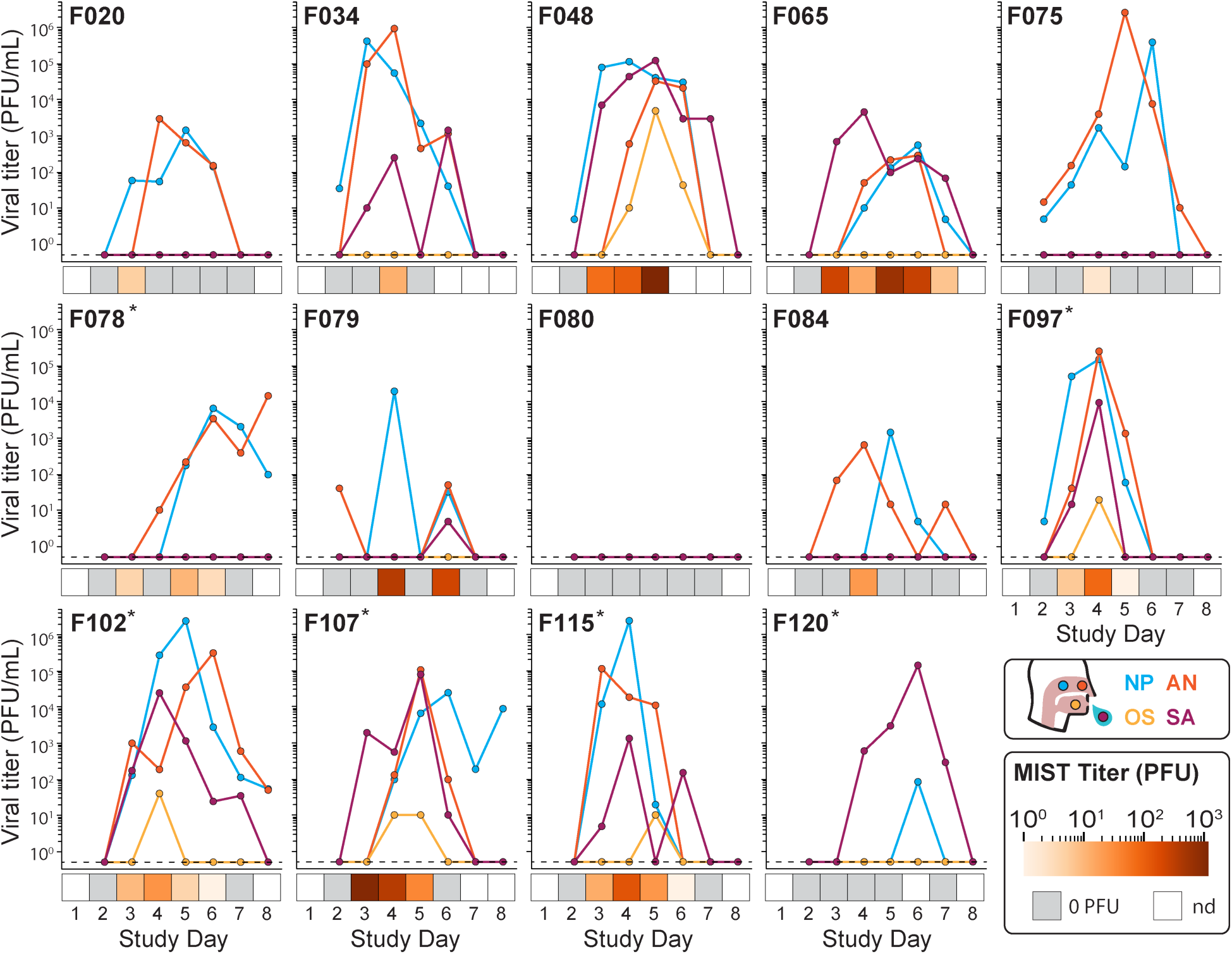
Viral load kinetics, spatial distribution, and extent of infectious viral expulsion differs across inoculated individuals. Viral loads observed in swabs from the nasopharyngeal site, the anterior nasal site, the oral cavity, and saliva. Each panel denotes a single participant. Identifiers with an asterisk highlight individuals inoculated via aerosol inhalation. Dashed horizontal lines denote the limit of detection of the plaque assay. Heat maps below the plots show the total number of plaque forming units (PFU) detected in MIST samples on a given day (including all plates for speaking/coughing and sneezing). Grey indicates an absence of PFU in MIST samples. White indicates MIST sampling was not done on that day (nd).

### Distinct genetic profiles characterize virus populations in different anatomical sites

The genetic composition of viral populations was assessed by whole genome sequencing of the challenge stock and all PFU positive samples. Samples were processed in duplicate from the RNA stage, yielding two independent sequencing datasets. Variants above a frequency of 1% were identified relative to a reference sequence for the influenza A/Perth/16/2009 (H3N2) virus. Variants were considered in subsequent analyses only if they were detected in both sequencing replicates of a given sample.

There were four minor variants present in the stock at relatively high frequency: Two in the NP gene segment that were of comparable frequency, suggesting genetic linkage (NP at N101D 0.31 and NP L136I at 0.30 frequency). Similarly, two in the PA gene segment showed evidence of genetic linkage (PA I211M at 0.35 and PA I228N at 0.37). In addition, several minor variants at 1-5% frequency were detected in the stock. Some, but not all, of these variants transferred to the participants (Supplementary Figure 10). Additional minor variants that were not observed in the stock could be identified in most of the clinical samples, with an average of 13.2 variants per sample. Most of these potentially de novo variants were observed at low frequencies (<0.15). Variant frequencies within a given participant were often variable over time, consistent with previous findings^17,29^ and likely reflecting stochastic dynamics (Fig. 3A). Variant composition and frequencies across sites on a given day were also variable, revealing spatial heterogeneity. In some examples, such as participant F065, viral genetic composition in saliva samples was highly dissimilar from that seen in anterior nasal samples. In most participants where multiple sites could be compared, however, populations within anterior nasal, nasopharyngeal and saliva samples showed more modest dissimilarity. These data are consistent with viral mixing across the sites sampled being either limited or sporadic. Nevertheless, as expected, dissimilarity of viral populations sampled from a given participant was typically lower than between participants (Supplementary Fig. 11).

**Figure 3.**
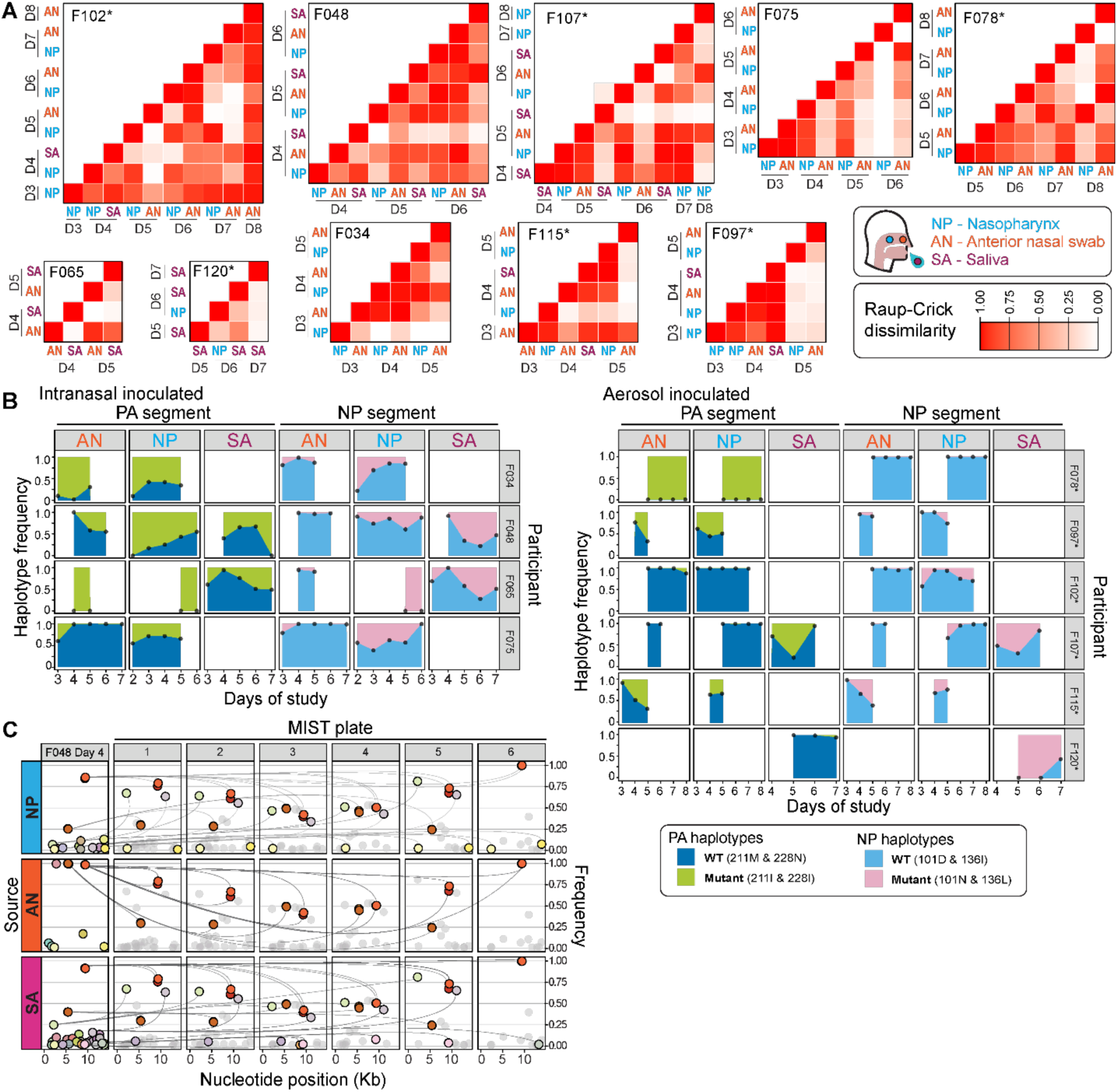
Viral populations differ across anatomical sites and are incorporated efficiently into infectious respiratory particles. A) Raup-Crick dissimilarity index comparing virus populations across sites and days evaluated. Values of 1 (white) show the highest dissimilarity in variant composition of the viral populations examined. B) Evolutionary trajectory of genotypes found in the viral PA and NP gene segments. Participants are identified at the right. All samples where the haplotypes were detected are included. C) Variants detected within participant F048 are related to those collected during coughing and sneezing in the MIST on the same day (study day 4). Each shared variant is assigned a unique color. Grey variants represent those unique to the MIST. Grey lines connect variants detected in the participant and the MIST. Y-axis on the right shows variant frequency. X-axis show nucleotide position in a concatenated version of the IAV genome.

The four nonsynonymous variants present in NP and PA at frequencies of 0.30–0.37 matched the reference strain and – likely due to their appreciable frequencies in the stock – were efficiently transferred to participants. Because the alternative NP and PA genotypes were observed across successive sampling days, they offered another opportunity to assess relationships between the sampled locations (Fig. 3B). Since the minor variants in NP and PA match the reference strain, we refer to them here as wildtype. Overall, the frequency of the major genotypes in NP and PA tended to decrease over time as they were replaced by the wildtype alternatives. These dynamics were often discordant across sites, however. For example, in F048, the wildtype NP genotype neared fixation in the anterior nasal site but remained the minor variant in saliva on three successive days. Similarly, in F065, the wildtype genotype in PA dominated in saliva but was not detected in either nasal location. Again, these observations point to constraints on mixing between oral and nasal viral populations.

### Virus populations within expulsions comprise multiple variants

The strategy of assessing infectious respiratory particles collected in the MIST by plaque assay allowed the genetic characterization of the expelled viruses through sequencing of plaque isolates. Because of the plaque-based culture, the composition of the viral population was likely skewed, however. Thus, although variant frequencies are obtained from next generation sequencing, we focused on the presence or absence of minor variants in MIST samples in our analyses. Thus, we compared the variants detected in up to 10 plaque picks from each plate of the MIST to those present in the bulk swab and saliva samples (Fig. 3C and Supplementary Fig. 12). Strikingly, many variants present in the within-host samples were also found in expelled particles, revealing robust transfer of viral diversity into the environment. Each variant was furthermore typically found across several MIST plates, indicating that a given variant was often present in multiple distinct infectious respiratory particles.

### Symptoms and infectious viral loads are positively associated with infectious respiratory particle production

With the goal of identifying likely anatomical sources and features of infection that may promote infectious respiratory particle production, we considered both viral sequencing data and parameters associated with infectious titers in MIST samples. First, with the rationale that source and sink populations would have similar genetic composition, the number of shared variants between distinct within-host samples and MIST samples collected on the same day was evaluated across participants (Fig. 4A). While shared variants were often detected in high numbers, no clear distinction among anatomical sites was apparent: the percentage of variants shared with MIST samples was comparable for saliva, nasopharyngeal, and anterior nasal samples (Fig. 4A). We then analyzed the relationships between infectious titers in MIST samples and in nasopharyngeal swabs or saliva (Fig. 4B, C). Both showed significant positive relationships in linear mixed effects models with levels of infectious virus in saliva (Akaike Information Criterion (AIC) = 181.06) being a better predictor of infectious respiratory particle expulsion than infectious virus in nasopharyngeal swabs (AIC = 193.37).

**Figure 4.**
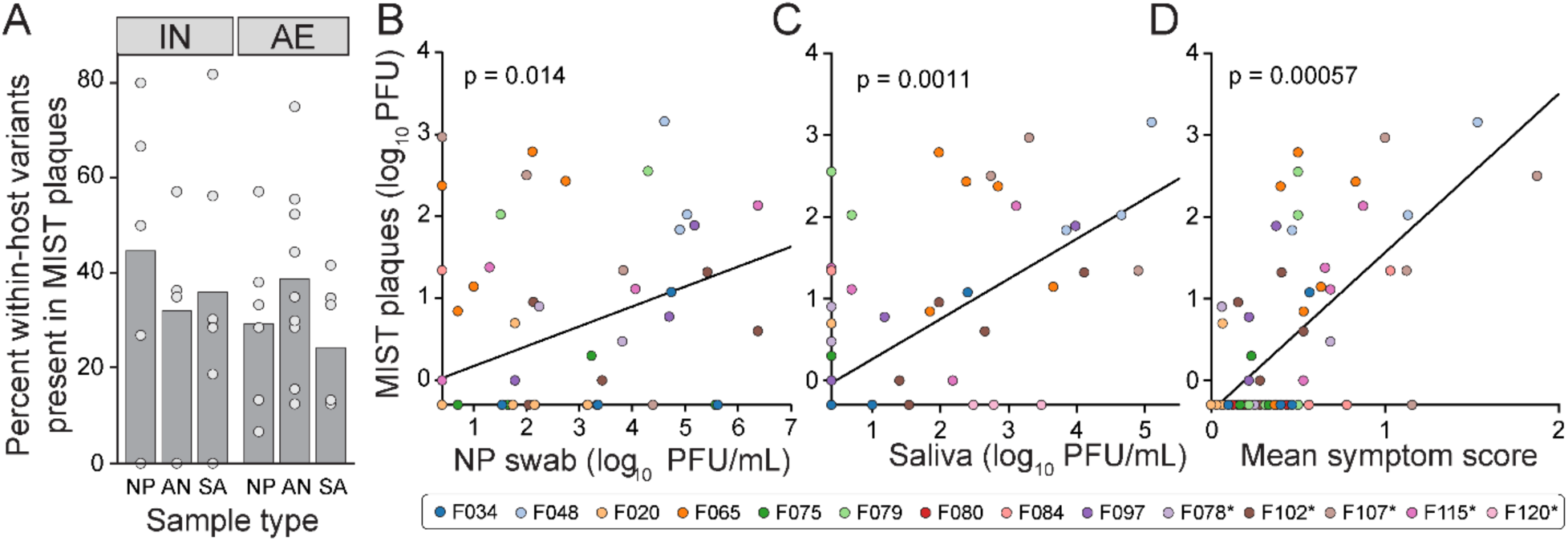
Positive correlates of infectious respiratory particle production include infectious viral loads in NP and saliva samples, and symptoms. In all panels, each data point represents a single participant on a single day. Infectious titers in MIST are summed across all plates. A) The percentage of all variants detected in nasopharyngeal (NP), anterior nasal (AN) or saliva (SA) samples that are also detected in plaques derived from the MIST plates. B–D) Linear mixed effects models of the relationship between infectious titers in the MIST and B) infectious virus in nasopharyngeal swabs, C) infectious virus in saliva, and D) mean total symptom scores. Samples are color coded by participant and are consistent in each sub-panel. Black lines indicate the regression lines from the models. p-values indicate the significance of the association between the fixed effect variable and the number of MIST plaques detected.

Finally, the relationship of specific categories of symptoms to infectious respiratory particle production was examined (Fig. 4D and Supplementary Fig. 4 and 13). The daily mean symptom score of all 32 Flu-Pro symptoms was found to have a significant association with infectious respiratory particle production. This outcome is likely driven by specific symptoms, with the categories including runny nose and sinus congestion, and headaches, body aches, and fatigue having the most significant associations with infectious respiratory particle production. These results support the notion that symptomatic cases of influenza may be more likely to seed infection in contacts.

## Discussion

Here we report quantification and genetic analysis of infectious respiratory particles produced by healthy adults infected with a seasonal IAV and relate these findings to within-host viral loads and sub-population compositions.

Examination of the infectivity of influenza virus in airborne particles has presented major technical challenges for decades. Instruments designed to differentiate particles by size are generally ineffective at preserving virus infectivity, while those designed to preserve infectivity have variable success^30,31^. The G-II sampler has consistently provided data on viral load in fine aerosols (<5 µm)^11^ but is not designed for analysis of particles larger than ∼5 µm^32^. An SKC liquid impinger has been successfully applied to detect infectious IAV in coughs and exhaled breath but does not fraction the collected aerosols by size and has poor efficiency for collection of particles >10-15 µm^14,33^. Isolation of infectious IAV from the breath of ferrets was achieved using the Influenza Virus Transmission Tunnel (IVTT)^27^. The IVTT leveraged gravitational settling of particles onto cultured cells as a sensor for infectious respiratory particles. Thus, the IVTT sampled larger particles and used direct viral culture to overcome the loss of infectious material. Our work built on this concept, with the addition of features to quantify respiratory particles and to capture fine infectious respiratory particles that do not settle within a short distance. Particles and infectious respiratory particles produced by infected humans were readily detected along the 97 cm length of the MIST apparatus, providing quantification of airborne viral dissemination within this range. Tests with synthetic aerosols suggest that particles settling within the MIST are mainly those >10 µm in diameter. While particles unable to settle in the MIST were captured using either a BioSpot VIVAS or a PTFE filter, infectious respiratory particle collection by these methods was low, likely due to both inefficiency of these devices and low amounts of infectious virus contained within small particles.

By combining the MIST technology with a controlled human infection model, we were able to relate the production of infectious respiratory particles to the size and genetic composition of viral populations within the nasal and oral cavities. The magnitude of infectious viral expulsions was positively associated with both saliva and nasal infectious viral loads, but this association was stronger for saliva. Conversely, infectious expulsions did not show a clear relationship with the total volume of particles deposited, suggesting that these particles originate from multiple anatomical sites with varying viral occupancy and therefore comprise a mixture of infectious and non-infectious particles. The spatial heterogeneity of viral population density within a host and the production of respiratory particles from multiple sites likely create variation in the source of infectious respiratory particles. Nevertheless, our data suggest that saliva should be considered as a potential vehicle for transmission and that caution is needed in using nasopharyngeal viral loads, in particular genome copy data, as a proxy for infectiousness. Viral variants detected within the nose and mouth were often present within expelled samples. The same variant could furthermore be seen at multiple sampling locations within the MIST, documenting repeated incorporation into respiratory particles. These observations indicate an efficient transfer of viral diversity from the host to the environment, indicating that tight genetic bottlenecks associated with transmission^17^ are unlikely to arise during expulsion.

The observed heterogeneity among participants in symptoms, viral load dynamics, viral spatial distributions and infectious respiratory particle production suggest features of human influenza that may contribute to superspreading dynamics^34^. Secondary attack rates of IAV within households range from ∼5% to ∼20%^35–38^. Together with occasional reporting of large outbreaks^39^, the variability seen in secondary attack rates suggests that IAV transmission is heterogeneous, as is well-documented for SARS-CoV-2 and SARS-CoV^40,41^. The drivers of transmission heterogeneity remain unclear^34^. The potential for extreme viral shedding by rare individuals was previously highlighted with the identification of a high emitter that generated viral RNA copy numbers in exhaled breath that were 100x greater than the median^15^. While elevated particle emissions, such as those associated with increased loudness of speech^42,43^, have been suggested to play a role, our data indicate that heightened emission of respiratory particles may not yield increased shedding of infectious virus into the air. Conversely, the relationships observed herein between infectious respiratory particle production and viral loads suggest that heterogeneity in sites of viral replication and shedding into the oral cavity may be important factors. The correlation of certain symptoms with MIST titers further suggests that variation in disease manifestations may contribute to heterogeneity in infectious respiratory particle production and onward transmission.

For most participants, our data show higher MIST titers recovered from coughing and sneezing than from speaking. These data do not, however, indicate which respiratory activities are more likely to drive transmission, since the number of coughs and sneezes and the duration of sampling from speaking were set arbitrarily. The voluntary coughs and sneezes examined may furthermore differ from natural expiratory events. The positive association seen herein between coughing symptoms and infectious respiratory particles production may be more informative of the importance of coughing for transmission. Indeed, related prior studies suggest a role for coughing. In one study examining naturally infected participants, comparable rates of infectious virus positivity were seen when particles <10-15 μm were sampled from exhaled breath and from voluntary coughs^14^. In another, quantities of viral RNA in particles <5 µm were increased when participants coughed unprompted during sampling^15^. Of note, different expiratory maneuvers would be expected to produce particles from different parts of the respiratory tract and the oral cavity. These differences could drive differing contributions to transmission and are also important to consider in interpreting the data from the MIST. For example, MIST titers may show stronger association with saliva viral loads than nasal viral loads owing to the sampling from ten coughs and only three sneezes.

It is important to consider additional limitations of our study. First, while our goal is to advance understanding of IAV transmission, this work focuses on only the first step of that complex process, expulsion of virus from an infected individual. Definition of the relationship between expulsion and onward transmission is needed in future research. Second, since the study’s sample size was small, certain host characteristics that modulate infectious respiratory particle expulsion may not have been represented. Finally, the participants studied were infected experimentally, with a relatively high viral dose and through routes that may not mimic natural infection. These features of the model could shape viral tissue distribution, symptoms and other features relevant for infectious respiratory particle production. While the doses and routes relevant in natural infection are likely to vary widely, their relationship with the controlled human infection models employed here is unclear.

In conclusion, we report the robust detection of infectious respiratory particles produced by influenza virus infected humans, including particles of large volume. Our data underscore marked person-to-person heterogeneity in infectious expulsions and show that expelled, infectious particles often carry within-host viral diversity, two features likely to shape transmission opportunity and viral evolution in real-world settings.

## Materials and Methods

### Ethics Statement

This study was conducted at the Emory University Hospital in Atlanta, Georgia between July and October 2023 under IRB protocol STUDY00000083 (clinicaltrials.gov identifier NCT05332899) and in accordance with the Declaration of Helsinki and Good Clinical Practice guidelines. A US Food and Drug Administration (FDA) investigational new drug designation (IND #19579) was obtained for the influenza A/Perth/16/2009 (H3N2) challenge virus produced in accordance with Good Manufacturing Practice (GMP) by Meridian Life Sciences (Memphis, TN) on behalf of hVIVO (London, United Kingdom) originally described in^22^. During the study no severe symptoms, no complications and no staff exposure incidents were observed.

### Cells and Viruses

Madin–Darby canine kidney (MDCK) cells were a gift from Dr. Daniel Perez, University of Georgia, Athens, GA. A seed stock of MDCK cells at passage 23 was amplified and maintained in Minimal Essential Medium (Gibco) supplemented with 10% fetal bovine serum (FBS) (Atlanta Biologicals) and Normocin (Invivogen). Cells were cultured at 37 °C and 5% CO_2_ in a humidified incubator. All cell lines were tested monthly for *Mycoplasma* contamination while in use and discarded if found to be positive. The influenza A/Perth/16/2009 (H3N2) challenge virus (hVIVO) was provided from the manufacturer in single dose aliquots at a concentration of 5.5 log_10_ median tissue culture infective dose (TCID_50_)/mL. The stock was produced under good manufacturing practice (GMP) conditions and was passaged 6 times in eggs. The sequence was verified to be similar to the wild type as described in^22^. The stock was characterized genetically by viral whole genome sequencing on an Illumina platform, revealing four consensus-level polymorphisms that differ from the reference (KJ609203 – KJ609210): PA I211M, PA I228N, NP N101D and NP L136I. Raw sequence data from the stock are available at SRA SAMN45722272 and SAMN45722273. For hemagglutination inhibition assays, a stock of this virus was made by passaging the challenge virus once in embryonated hen’s eggs.

### Participant Recruitment and Study Design

This study was open to healthy persons aged 18 to 49 years^24^. Volunteers were initially screened for hemagglutination inhibition (HAI) titers against influenza A/Perth/16/2009 (H3N2) within 60 days of study enrollment. Hemagglutination inhibition assays were performed as described previously^24^. Briefly, serum was treated with receptor-destroying enzyme (Hardy Diagnostics, Santa Maria, CA), heat inactivated at 56 °C, serially diluted, mixed with 4 HA units of virus, incubated at room temperature for 15 minutes, mixed with an equal volume of 0.75% turkey red blood cells (Lampire Biological, Pipersville, PA), and incubated at room temperature for 40 minutes. HAI titers were determined by the inverse of the highest sera dilution that inhibited hemagglutination. Those with HAI titers less than or equal to 40 were subsequently screened to assess medical histories and physical health within 30 days of study enrollment. A full list of eligibility requirements are listed in the supplementary information. Twenty-four volunteers who met all eligibility criteria, demonstrated knowledge and comprehension of the study, and provided written informed consent were selected to participate in this study.

The twenty-four participants were enrolled across ten cohorts occurring between July 2023 and March 2025. Participants were admitted to Emory University Hospital on study day 0 and quarantined in single-occupancy hospital rooms for eight nights. Intranasal inoculation with the challenge virus occurred on study day 1 using a Mucosal Atomization Device (MAD) (Teleflex, Wayne, PA) to deliver 0.5 mL into each nostril for a total dose of 5.5 log_10_ median TCID_50_ in a 1 mL volume^22,24^. The Mucosal Atomization Device is designed to produce particles with diameters ranging from 30 to 100 μm, based on manufacturer specifications^47^. A study utilized laser diffraction and a cascade impactor to validate the specifications and measured a volume median aerodynamic diameter in the range of 80-100 µm^48^. The device has been used with similar inoculum volumes (1 mL) and similar doses in other human challenge studies^49,50^. Participants adopted a supine position during virus administration and for 30 seconds after. Inoculation via aerosol inhalation was performed as described in^25^. Briefly, aerosols containing influenza viruses were produced using a flow-focused mono-dispersed aerosol generator (FMAG) or jet nebulizer into a fit test hood where participants breathed naturally for 10-20 min. Participant health was monitored regularly during the quarantine period through daily vital sign collection and symptoms were reported on study days 2-8 using an inFLUenza Patient-Reported Outcome (FLU-PRO^©^) survey^26^. Participants were discharged from the hospital on study day 8. Oseltamivir phosphate was provided at the time of discharge unless negative clinical tests for influenza RNA were obtained on both study days 7 and 8 (see sample collection section).

### Sample collection

Upper respiratory tract samples were collected from participants once daily on study days 0 and 2–8. Nasopharyngeal (NP) swabs were collected by swabbing the nasopharynx with a flexible mini-tipped flocked swab. Anterior nasal (AN) swabs were collected by swabbing the anterior nares, 1-2 cm deep in each nostril, in a circular motion with a flexible minitipped flocked swab. Oral swabs (OS) were collected by swabbing the inside of both cheeks and underneath the tongue for 10-15 seconds at each site using a regular tipped flocked swab. All swabs were placed into tubes containing universal viral transport medium (BD, Franklin Lakes, NJ). NP swabs were immediately tested at the Emory University Hospital microbiology laboratory using the Xpert Xpress CoV-2/Flu/RSV plus test (Cepheid, Sunnyvale, CA), a multiplexed real-time PCR which detects influenza virus RNA using two distinct primer sets (Supplemental Figure 1). All swabs were refrigerated for less than 4 h prior to being vortexed vigorously for 10 seconds, aliquoted, and frozen at –80 °C. Approximately 3 mL of Saliva was self-collected by participants spitting into collection cups. Saliva was refrigerated for less than 4 h prior to being aliquoted and frozen at –80 °C.

### Titration of infectious virus

Infectious virus was quantified by plaque assay on confluent MDCK monolayers. Samples were serially diluted in 1X PBS (Corning) and 200 µl of each dilution added to 6-well plates after removing culture medium and washing with 1X PBS. Cells were incubated for 1 hour at 37°C, with occasional rocking. After incubation, the inoculum was removed, cells were washed with 1X PBS, and overlayed with plaque assay medium (2X Minimal Essential Media (Gibco) supplemented with 4 mM L-glutamine (Gibco), 30 mM HEPES buffer (Corning), 2X Penicillin-Streptomycin Solution (Corning), 0.3% sodium bicarbonate (Corning), 0.42% bovine serum albumin (BSA) (Sigma-Aldrich), 0.01% diethylaminoethyl (DEAE)-dextran (MP Biomedicals), 1 µg/mL tosyl phenylalanyl chloromethyl ketone (TPCK)-treated trypsin (Sigma-Aldrich) and 0.7% Oxoid agar (Oxoid)). Plaques were counted by visual inspection under oblique lighting after a two-day incubation at 37°C.

### Modular Influenza Sampling Tunnel (MIST) design

The MIST was designed to sample infectious respiratory particles of all sizes while maintaining a compact footprint. A participant positions their nose and mouth in the opening of a thin, rubber sheet that covers a circular portal at one end. At the opposite end, an AeroTrak particle counter (TSI Inc., Shoreview, MN) running at 2.8 L/min and a BioSpot VIVAS sampler (Aerosol Devices, Fort Collins, CO) or an open-face filter collecting at 8.0 L/min continuously sample air from the MIST, for a total flow rate of 10.8 L/min. Virus in ambient air should be negligible because we operated five portable HEPA air cleaners in the room during sampling and, in a separate study^24^, we have not detected virus in air samples collected in the rooms of infected participants. In between each sample collection, we reset the MIST by replacing the culture plates and water-sensitive paper after cleaning twice with wipes containing Clorox® (0.145% n-Alkyl (60% C14, 30% C16, 5% C12, 5% C18) dimethyl benzyl ammonium chloride, 0.145% n-Alkyl (68% C12, 32% C14) dimethyl ethyl benzyl ammonium chloride) to remove virus deposited in the tunnel. Additionally, we ran a small, portable fan inside the MIST for 5 minutes to remove lingering aerosols generated during each sampling session.

*Analysis of settling particles.* Particle sizes which settle within the MIST can be detected along the length of the tunnel using strips of water-sensitive paper (Innoquest, Inc.) placed in between the culture plates; the yellow paper turns blue where droplets fall onto it. Image processing software (ImageJ v1.54d) was used to quantify the number and size of the blue spots (Supplementary Fig.14). The size of deposited droplets on the water-sensitive paper was adjusted to estimate their original volume using a published spread factor for respiratory expulsions^51,52^. The minimum particle size that we can be detected using the water-sensitive strips was approximately 20 µm in diameter. This limit was determined by comparing the minimum pixel size measured using the ImageJ software to the total pixel area of the strip (3 inch^2^).

The cell culture plates in the MIST are designed to directly culture infectious virus expelled into the air within infectious respiratory particles. Plates (L:127.76 mm x W:85.47 mm x H:32 mm) at six positions were included to define infectious content of particles with different size distributions. Plates were pre-seeded with MDCK monolayers. Following sampling, plates were overlaid with plaque assay medium to ensure that each infectious unit formed a single plaque. After 48 h of incubation, the number of plaques were quantified.

*Analysis of non-settling particles within MIST.* Air sampling devices at the distal end of the MIST capture small aerosol particles (up to 20 µm in diameter) that remain airborne for extended periods. An AeroTrak was used to count aerosol particles by size. A BioSpot VIVAS was used to collect aerosol particles at a flow rate of 8 L/min into PBS containing 0.2 M sucrose and 0.5% bovine serum albumin for the purpose of quantifying viral material for participants F020, F034, F048, F065, F075, F079, F080, and F084. In place of the BioSpot VIVAS, an open-face 37 mm, 3.0 μm pore polytetrafluoroethylene (PTFE) filter (Millipore) was used to collect air at 8 L / min using a Gilian Gilair 12 pump (Sensidyne, St. Petersburg, FL) for participants F078, F097, F102, F107, F115, and F120. This filter was then placed in a 1 mL of universal viral transport media and vortexed for one minute to elute the material collected. Aerosol samples derived from the BioSpot VIVAS or the PTFE filter were stored at –80°C and then tested by plaque assay for detection of infectious virus and qRT^5321^¾. Samples identified as containing influenza genomes by one-step quantitative RT-PCR were retested using a second independent nucleic acid extraction. Both the first and second extractions were tested by qRT-PCR in technical duplicate. Ct values were converted to genome copy equivalents using a standard curve generated by testing serial dilutions of an *in vitro* transcribed A/Perth/16/2009 (H3N2) M gene segment.

### Characterization of particle deposition in aerosol sampling devices

The sampling capabilities of the MIST were validated by generating synthetic aerosols of viral preparations with known infectivity. Influenza A/Perth/16/2009 virus was diluted in PBS and aerosols were produced with three aerosol generation devices: a jet nebulizer designed to generate particles with 0.5–2 µm diameter, a flow-focusing monodisperse aerosol generator (FMAG) set to produce particles with 10 µm diameter, and a mucosal atomization device (MAD) designed to produce particles with 30-100 µm diameter (per manufacture’s technical details). Three trials were run with each aerosol generation method. For each, the recovery of infectious virus and viral genomes on the cell culture plates of the MIST and the PTFE membrane sampling from the distal end of the MIST was evaluated. In addition, the collection of total particles on water sensitive paper strips and by the Aerotrak sampling from the distal end of the MIST were evaluated. The efficiency of collection of infectious respiratory particles using the BioSpot VIVAS and a PTFE membrane was further examined separately from the MIST tunnel. Each was used to collect infectious aerosols delivered into a 4 L chamber by the jet nebulizer or the FMAG, with collection occurring a few inches from the aerosol generation source.

### Modeling droplet deposition in the MIST

Particles undergo evaporation after being expelled into the tunnel, leading to a reduction in size and increased suspension times. We modeled particle transport within the MIST using a system of non-linear differential equations. The equilibrium size of the evaporating droplet (D_p_) was determined using the following mass and heat transfer equations:

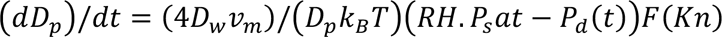

Where

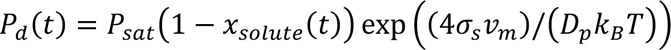

Where D_p_ is the diameter of the droplet, D_w_ is the diffusivity of water vapor, v_m_ is the molecular volume of water, k_B_ is the Boltzmann’s constant, T is the temperature, RH is the ambient relative humidity, P_sat_ is the saturation vapor pressure of water, P_d_ is the vapor pressure of water at the droplet’s surface, x_solute_ is the mole fraction of salts in the droplet, σ_s_ is the surface tension of water, and F is the Fuchs-Sutugin correction factor calculated as:

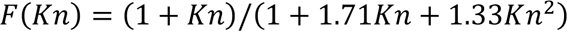

Where Kn is the Knudsen number (which is the ratio of the mean free path of the vapor molecules to the radius of the particle). The correction factor F accounts for change in transport dynamics at the droplet surface of small particles.

Particle transport in the horizontal direction is the sum of the expiration velocity and the velocity of the ambient air. The ambient air velocity was assumed to be 0.34 cm/s, estimated as the ratio of the total volumetric flowrate of sampling devices to the cross-sectional area of the MIST. The expression used for the cough front velocity (u_cough_) was based on Particle Image Velocimetry measurements performed by Savory et al.:

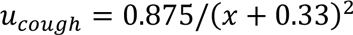

Where x is the horizontal distance from the participant. The speaking front velocity was assumed to be u_cough_/3^54^. Vertical settling of the human-expelled particles was estimated by solving the following momentum balance equation:

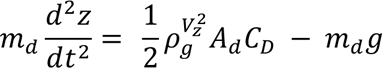

Where z is the vertical settling distance, m_d_ is the mass of the droplet, ρ_g_ is the density of air, V_z_ is the vertical velocity of the particle, A_d_ is the cross-sectional area of the particle, C_D_ is the drag coefficient based on the Reynold’s number and g is gravitational acceleration.

### MIST Sampling

Inoculated individuals confirmed positive for infection by qPCR were sampled using the MIST while speaking and coughing and sneezing. For the first cohort examined (which included participants F034 and F048), sampling was performed daily on Study Days 2–5. Collections were extended to Study Days 2–7 for subsequent cohorts since shedding remained robust at Study Day 5. Occasionally, sampling sessions were missed if a participant did not feel well enough to perform the task. Before sample collection, six single-well plates (ThermoScientific and CELLTREAT) with a confluent monolayer of MDCK cells containing 4.0 mL of 1X PBS and 2X Antibiotic-Antimycotic (Gibco) were placed in the tunnel with water-sensitive paper strips (1”x 3”) (Innoquest, Inc.) between each plate. Plates and water-sensitive paper were replaced between different expiratory activities. During speaking activities, participants performed a series of vocal exercises at an audible volume for 10 minutes. During the first 5 minutes, participants sang nursery rhymes and counted to 100. During the second 5 minutes, participants spelled words. The pump was turned off after the 10 minutes of activity, so some residual particles within the chamber may have been missed. To sample coughing and sneezing, participants were asked to simulate ten coughs followed by three sneezes. Both the AeroTrack and BioSpot VIVAS or Gilair 12 pump operated continuously during these activities and remained on for a total of 10 minutes. After sample collection, plates were incubated at room temperature for 40 min to 2 h, followed by overlay with plaque assay media. Overlayed plates were incubated for 48 h at 37°C, 5% CO_2_. As described above, the MIST was cleaned between participants.

### Plaque quantification and processing for whole-genome sequencing from MIST

Plaques on MIST plates were counted manually through direct visualization of unfixed monolayers under oblique lighting. The maximum limit of detection per plate was 250 PFU; quantities greater than this were recorded as ‘too many to count’ and included in the datasets analyzed herein as 250 PFU. Agar plugs were picked using a 1 mL serological pipette (VWR), followed by ejection into 160 µL of 1X PBS, and then stored at −80°C. Up to ten plaque picks from each plate were pooled: If a plate contained fewer than ten plaques, all were analyzed; if a plate contained more than ten plaques, ten plaques were analyzed. RNA was extracted from agar plugs using the QiaAmp Viral RNA kit (QIAGEN) with the following modifications to the manufacturer’s protocol: carrier RNA was not used, and the RNA was eluted in 30 µL of nuclease-free water.

### Whole-genome sequencing

Nasopharyngeal samples that were positive by qPCR or plaque assay, and anterior nasal, oral, and saliva samples that were positive for infectious virus were selected for whole-genome sequencing. Single plaques were not isolated for genomic sequencing. The viral genome was converted to cDNA and amplified using the Uni/Inf primer set in a multi-segment-RT PCR (M-RTPCR) as previously described^55^. DNA was generated from each RNA extract in duplicate and processed through all downstream steps, yielding two independent sequencing data sets per sample. Amplification products were visualized by electrophoresis in a 0.8% agarose gel (UltraPure™ Agarose, Invitrogen). Amplicons were purified using 0.45x AMPure XP beads (Beckman Coulter) and genomic material was quantified using the Qubit dsDNA HS Assay Kit (Invitrogen) in the Qubit 4 Fluorometer (Invitrogen). Library preparation (Nextera XT, Illumina), sequencing (NovaSeq 6000, 2×100 bp paired end reads, Illumina), and demultiplexing was performed at the Emory National Primate Research Center Genomics Core.

### Variant analysis

To increase sensitivity for detecting true variants, samples were sequenced in replicates. For the analysis, only samples with complete genome and gene segments with a mean depth equal or above 400 were considered. Variant analysis from samples replicates meeting the quality criteria were merged. Variants detected in both replicates were considered for the analysis whereas those that were unique were discarded.

Variant analysis was conducted using a custom pipeline^56^. Cutadapt was used to remove adapters followed by mapping of the reads to the reference genome using BWA-MEM^57^. Samtools was used to fix mate information, and LoFreq Viterbi provided probabilistic realignment to correct mapping errors. Reads were subsequently sorted with Samtools, and indel qualities were assigned using LoFreq indelqual. Base and indel alignment qualities were incorporated with LoFreq alnqual. Variant calling was executed with LoFreq, generating a VCF file. Custom scripts filtered and formatted this VCF to retrieve allele frequencies and coverage statistics, retaining only variants with a frequency of 0.01 or higher and a coverage of at least 400. SNPdat^58^ was then used to identify synonymous and nonsynonymous mutations.

### Raup-Crick dissimilarity indexes

To analyze genetic relationships between populations we used Raup-Crick dissimilarity index^59^. The Raup-Crick index, which considers the species shared between two populations, was applied with species represented by unique variants present in the genome. This index was chosen due to its robustness in calculating dissimilarity for unevenly sampled populations, ensuring that the calculated dissimilarities are accurate even when some samples have more or fewer available data points than others. The Raup-Crick index is sensitive to the total number of species—represented as variant sites—compared across samples; therefore, slight variability in dissimilarity calculations may occur when the total number of samples being compared varies. Calculation of Raup-Crick dissimilarity indexes was done using the vegan package version 2.6-4 in R.

### Statistical analyses

Relationships between viral titers, symptoms, expelled droplet volumes, and the amount of expelled infectious virus were assessed using linear mixed effects variable slope models allowing for each participant or participant and day (for expelled droplet volumes) to have a unique intercept and slope in R (v4.5.1) using the lme4 (v1.1.37) and lmerTest (v3.1.3) packages. Samples in which no plaques were detected were given values of half of the limit of detection (5 PFU/2 = 2.5 PFU for viral titers, and 1 PFU/2 = 0.5 PFU for expelled infectious virus). Likewise, water sensitive strips with undetectable droplet volumes were assigned a value of half the limit of detection (1×10^−3^ µL / 2 = 5×10^−4^ µL). Numerical conversions of symptom survey responses were assumed to be linearly spaced for the purpose of these analyses. The data from these models was plotted using GraphPad Prism version 10.0.3 (GraphPad Software, Boston, MA). Paired t-test and linear regressions for aerosol data were performed using the ttest_rel and linregress functions in the SciPy package. The Raup-Crick dissimilarity index from the viral sequencing data was calculated in RStudio (v4.1.3) using the vegan package (v2.6.4). The “r1” null model was applied, with the number of simulations set to 1000. Normality was assessed using the shapiro.test method in RStudio. Pairwise statistical comparisons were performed using the wilcox_test method.

## Data Availability

All data produced in the present study are available upon reasonable request to the authors. Posting of data in online repositories is in progress at the time of submission.

## Acknowledgements

We thank the individuals that participated in this study. We also thank the nursing staff at the Emory University Hospital and members of the Hope Clinic team for their assistance during this study. In particular: Jianguo Xu (challenge stock preparation); Lisa Harewood and Evan Gutter (regulatory support); Cecilia Zhang, Vanessa Buster, Kareem Bechnak, and Sarah Bechnak (clinical research coordination), Dean Kleinhenz (finances), Alexandra Koumanelis (recruitment), Eduardo Monarrez (quality management), Brandi Johnson and the lab staff (processing of samples). We thank the Georgia CTSA for logistical and clinical support. We thank Matthew Gaddy, Julie Nguyen, and Kaitlyn Bushfield for assistance with sample testing and processing and Alexandra Longest and Vikas Goel for assistance with analysis of aerosol particle data. This work was funded by Flu Lab, NIAID Centers of Excellence for Influenza Research and Response (CEIRR), contract number 75N93021C00017, and internal Emory University funds awarded to NGR. NVM is supported by 1F31AI186480-01. Next generation sequencing services were provided by the Emory NPRC Genomics Core which is supported in part by NIH P51 OD011132. Sequencing data was acquired on an Illumina NovaSeq 6000 funded by NIH S10 OD026799.

## Author Contributions

Conceptualization: NVM, NS, NGR, LCM, ACL, SSL

Clinical trial design: NGR, SSL

Clinical study implementation: NGR, MDP, HM, JT, VS, RT, DG, CSK

Data collection: NVM, NS, MDP, KP, SD, AS, MJS, MNV, AJC, KB, VR, JP, SSL

Instrument design and validation: NS, AJP, MDP, AS, LCM

Data analysis: NVM, NS, LMF, MDP, DV

Preparation of initial manuscript draft: NVM, NS, MDP, DV, ACL, LCM, SSL

Review and editing of manuscript: All authors

Funding acquisition: NGR, ACL, LCM, SSL

