## Supplementary Figures for "Controlled human influenza infection reveals heterogeneous expulsion of infectious virus into air"

**Table of Contents**


Supplementary Information. Subject eligibility criteria

Supplementary Figure 1. Study consort diagram.

Supplementary Figure 2. Molecular detection of viral genomes in nasopharyngeal swabs.

Supplementary Figure 3. Thirty-two symptoms in the FLU-PRO© questionnaire were divided in nine symptom categories.

Supplementary Figure 4. Duration and intensity of symptoms varied across individuals

Supplementary Figure 5. Modular influenza sampling tunnel (MIST)

Supplementary Figure 6. Characterization of particle capture in the MIST.

Supplementary Figure 7. Respiratory particles detected in MIST

Supplementary Figure 8. Lack of a clear relationship between expelled respiratory particles and plaque forming units (PFU)

Supplementary Figure 9. Carry-over of variants from inoculum to participants

Supplementary Figure 10. Viral populations sampled across time and locations within one participant show lower dissimilarity than those sampled from unrelated participants

Supplementary Figure 11. Genetic composition of viral populations in infectious respiratory particles produced during coughing and sneezing

Supplementary Figure 12. Relationship between symptoms and expulsion of infectious virus

Supplementary Figure 13. Analysis of water-sensitive strips

**Supplementary Information**

**Subject eligibility criteria**

Subjects must meet all of the following criteria:

1. Provide written informed consent prior to initiation of any study procedures.
2. Are able to understand and comply with all planned study procedures.
3. Healthy males and non-pregnant, non-breast-feeding females aged ≥18 and ≤49 years of age inclusive at enrollment.
4. Women of childbearing potential must be practicing abstinence or using an acceptable method of birth control for at least 30 days prior to enrollment through the duration of the trial. Male subjects must agree not to father a child for the duration of the trial.
    a. A woman is considered of childbearing potential unless post-menopausal (absence of menses for ≥1 year) or surgically sterilized (tubal ligation/salpingectomy, bilateral oophorectomy or hysterectomy).
    b. Acceptable contraception methods for women include but are not limited to: sexual abstinence from intercourse with men, monogamous relationship with vasectomized partner, barrier methods such as condoms or diaphragms with spermicide or foam, effective devices (IUDs, NuvaRing®) or licensed hormonal products such as implants, injectables or oral contraceptives.
5. Women of childbearing potential must have a negative serum or urine pregnancy test at screening and negative urine pregnancy test within 24 hours prior to challenge.
6. Are in good general health, as determined by the study investigator within 30 days of challenge and do not have any of the following conditions:
    a. Chronic pulmonary disease (e.g., asthma, emphysema).
    b. Chronic cardiovascular disease (e.g., cardiomyopathy, congestive heart failure, cardiac surgery, ischemic heart disease, known anatomic defects).
    c. Chronic medical conditions requiring close medical follow-up or hospitalization during the past 5 years (e.g., diabetes mellitus, renal dysfunction, hemoglobinopathies).
    d. Immunosuppression or ongoing malignancy or history of malignancy (excluding nonmelanotic skin cancer in remission without treatment for more than 5 years)
    e. Neurological and neurodevelopmental conditions (e.g., cerebral palsy, epilepsy, stroke, seizures).
    f. History of postinfectious or postvaccine neurological sequelae.
    g. Autoimmune, inflammatory, vasculitic or rheumatic disease, including but not limited to systemic lupus erythematosus, polymyalgia rheumatica, rheumatoid arthritis or scleroderma.
7. Demonstrate knowledge and comprehension of the study by scoring ≥70% on a quiz of the study protocol and policies.
8. Agrees to not use cigarettes, e-cigarettes, marijuana, or other tobacco products during the quarantine period.
9. Agrees to not use prescription or over-the-counter medications that could impact influenza challenge efficacy or symptoms (including oseltamivir, zanamivir, peramivir, baloxavir marboxil, amantadine and rimantadine, aspirin, intranasal steroids, acetaminophen, decongestants, antihistamines, and other NSAIDs), within 14 days prior to quarantine and through the quarantine period, unless approved by the investigator.

**Subject Exclusion Criteria**

Subjects eligible to participate shall not meet any of the following exclusion criteria:

1. Have household contact with or have daily contact with: ‘
    a. Children under 5 years of age.
    b. Children and/or teenagers who are receiving long-term aspirin therapy.
    c. Women who are pregnant or who are trying to become pregnant.
    d. Persons older than 65 years of age.
    e. Persons of any age with significant chronic medical conditions such as:
        i. Chronic pulmonary disease (e.g., asthma).
        ii. Chronic cardiovascular disease (e.g., cardiomyopathy, congestive heart failure, cardiac surgery, ischemic heart disease, known anatomic defects).
        iii. Contacts who required medical follow-up or hospitalization during the past 5 years because of chronic metabolic disease (e.g., diabetes mellitus, renal dysfunction, hemoglobinopathies).
        iv. Immunosuppression or cancer.
        v. Neurological and neurodevelopmental conditions (e.g., cerebral palsy, epilepsy, stroke, seizures).
2. Are healthcare workers with patient contact in the 2 weeks after influenza challenge.
3. Plan to be living in a confined environment (e.g. ship, camp, or dormitory) within 2 weeks after receiving the challenge strain.
4. Females who are pregnant or plan to become pregnant at any time between the Screening Visit through the duration of the trial.
5. Females who are breastfeeding or plan to breastfeed at any given time throughout the study.
6. Have a body mass index (BMI) less than or equal to 18.5 and greater than or equal to 35.
7. Smoke more than 4 cigarettes, e-cigarettes, marijuana, or other tobacco products on weekly basis within 60 days prior to challenge.
8. Have moderate or severe illness and/or an oral temperature ≥100°F and/or diarrhea or vomiting within seven days prior to challenge.
9. Have a pulse rate less than 55 beats per minute (bpm) or >100 bpm. If heart rate is
    a. <55 bpm and the investigator determines that this is not clinically significant (e.g., athletes) and heart rate increases >55 bpm on moderate exercise (two flights of stairs), subject will not be excluded.
10. Have a systolic blood pressure less than 90 mmHg or greater than 140 mmHg on two separate measurements (screening and pre-challenge).
11. Have a diastolic blood pressure less than 50 mmHg or greater than 90 mmHg on two separate measurements (screening and pre-challenge).
12. Have long-term (≥2 weeks) use of high-dose oral (≥20 mg per day prednisone or equivalent) or parenteral glucocorticoids, or high-dose inhaled steroids for greater than 7 days in the last 3 months.
13. Have an active HIV, hepatitis B, or hepatitis C infection.
14. Have screening laboratory test results (white blood cells (WBCs), absolute neutrophil count (ANC), hemoglobin (Hgb), platelets) that are outside the laboratory reported normal values and deemed clinically significant by the study investigator.
15. Have a serum creatinine greater than 1.1 x upper limit of normal (ULN).
16. Have an alanine aminotransferase (ALT) greater than 1.1 x ULN.
17. Have abnormal findings on screening electrocardiogram deemed clinically significant by study investigator.

18. Have abnormal findings on screening chest X-ray deemed clinically significant by study investigator.
19. Have ongoing drug abuse/dependence (including alcohol), or a history of these issues within 5 years of enrollment.
20. Have positive urine/serum test for drugs of abuse (i.e., amphetamines, cocaine, benzodiazepines, opiates, or metabolites) but not tetrahydrocannabinol (THC) or metabolites).
21. Have any medical, psychiatric, occupational, or behavioral problems that could make it difficult for the subject to comply with the protocol as determined by the investigator.
22. Have received experimental products within 30 days before study entry or plan to receive experimental products at any time during the study.
23. Plans to enroll in another clinical trial that could interfere with safety assessment of the investigational product at any time during the study period, including study interventions such as drugs, biologics or devices.
24. Plan to donate blood during the course of the study.
25. Have received a live vaccine within 30 days before study entry or plan to receive a live vaccine prior to Day 31 of the challenge.
26. Have received an inactivated vaccine within 14 days before study entry or plan to receive an inactivated vaccine prior to Day 14 of the challenge.
27. Have received parenteral immunoglobulin or blood products within 3 months of the study start, or plan to receive parenteral immunoglobulin or blood products for the duration of the study.
28. Have a known close contact with anyone known to have influenza in the past 7 days prior to screening or challenge.
29. Have a known history of allergy to anti-influenza drugs, more than 2 classes of antibiotics or severe egg allergy.
30. Have any condition that, in the judgment of the study investigator, is a contraindication to protocol participation or impairs the subject’s ability to give informed consent.
31. Have a BIOFIRE® FILMARRAY® respiratory panel that identifies any pathogen on the day of admission.
32. Clinically significant abnormality as deemed by the study investigator on pulmonary function testing and/or spirometry at screening visit (aerosol inoculation group only).

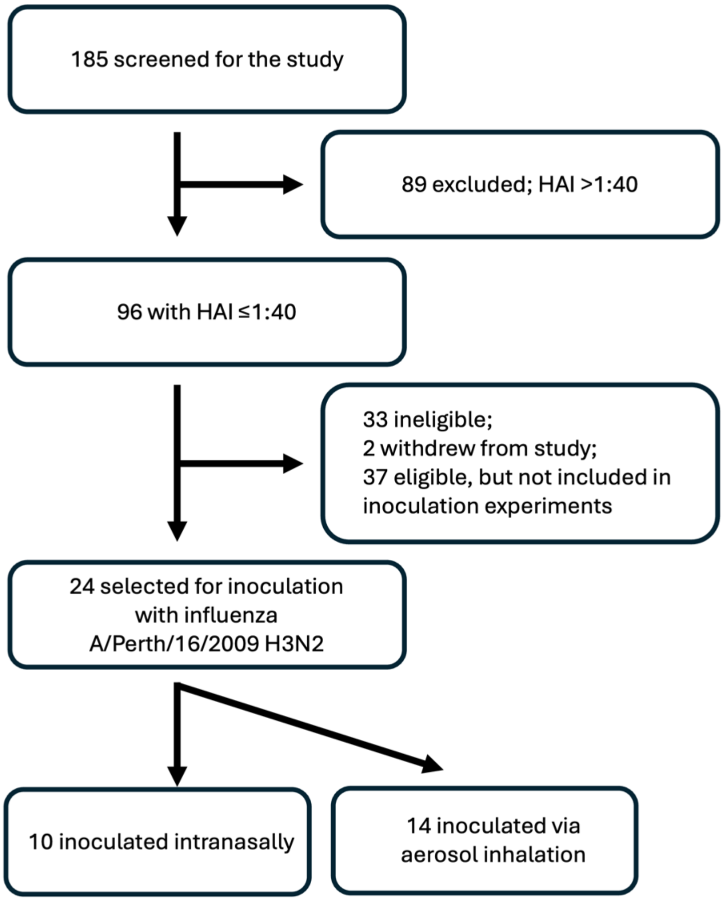


**Supplementary Figure 1. Consort diagram summarizing the recruiting and enrollment in the study.**

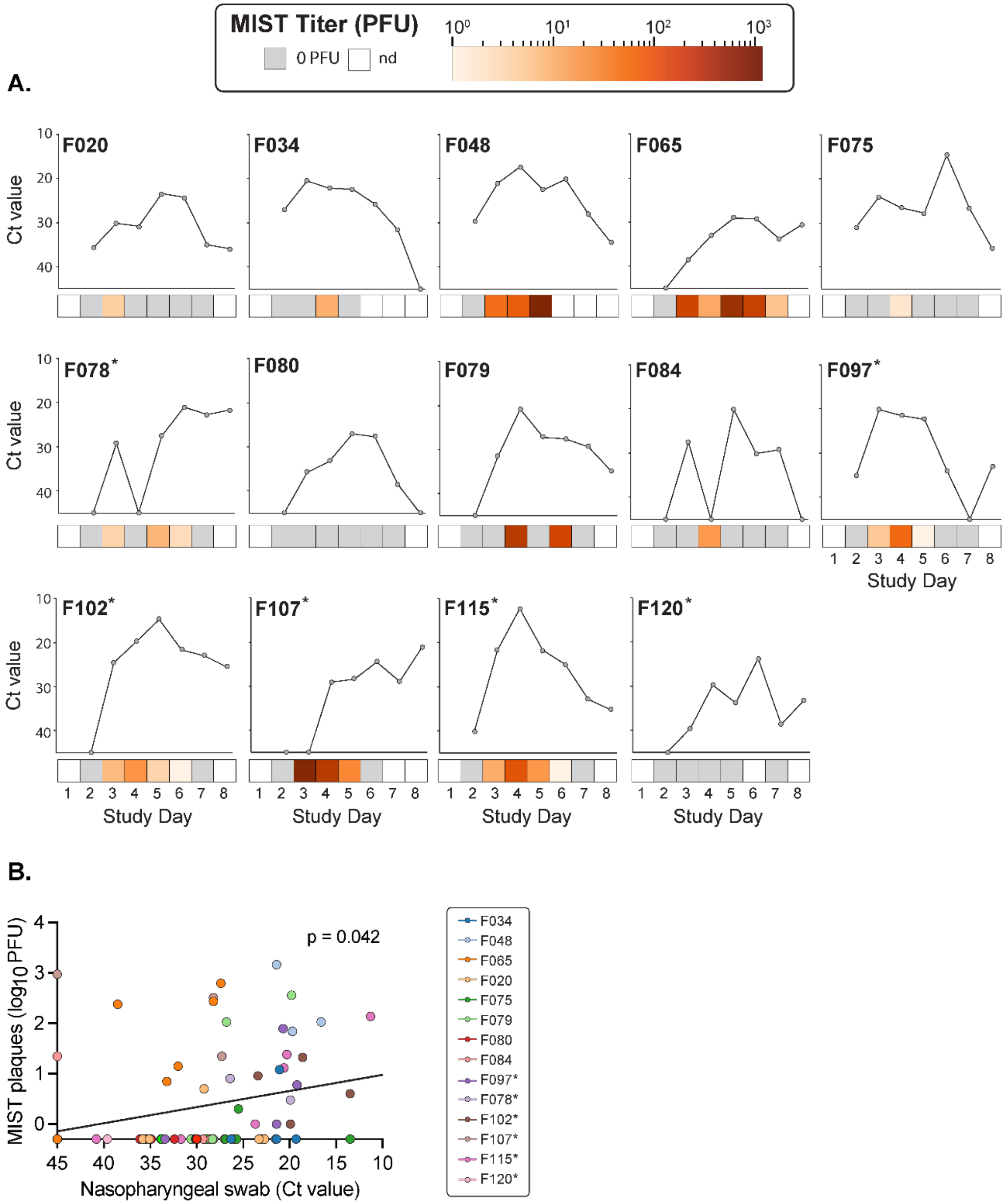


**Supplementary Figure 2. Molecular detection of viral genomes in nasopharyngeal swabs.** A) Data show the mean of the Ct values from two influenza virus genetic targets. One nasopharyngeal swab per participant was collected daily. Participants were inoculated on study day 1. Plaque forming units (PFUs) detected in MIST are shown with a heat map underneath each plot. Titers displayed are the sum of PFUs detected on all twelve plates in MIST during a given sampling day (inclusive of speaking and coughing and sneezing). Participant identification numbers are shown in the upper left of each facet. B) A linear mixed effects model of the relationship between infectious titers in the MIST and the Ct value from diagnostic testing of nasopharyngeal swab samples. Regression is shown using a black line. p-values indicate the significance of the association between the two variables. Each participant is indicated using a different color.

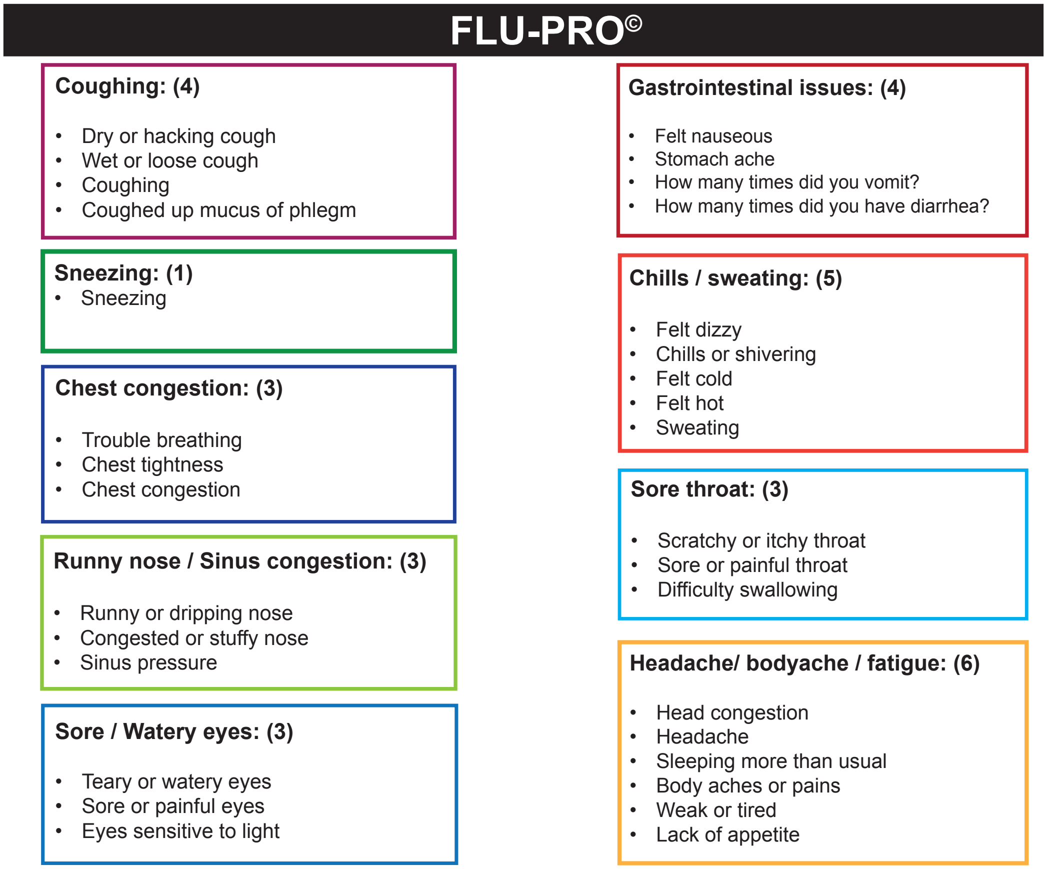


**Supplementary Figure 3. Thirty-two symptoms in the FLU-PRO© questionnaire were divided in nine symptom categories.** Each box includes a symptom category. The number of symptoms per category are in parentheses.

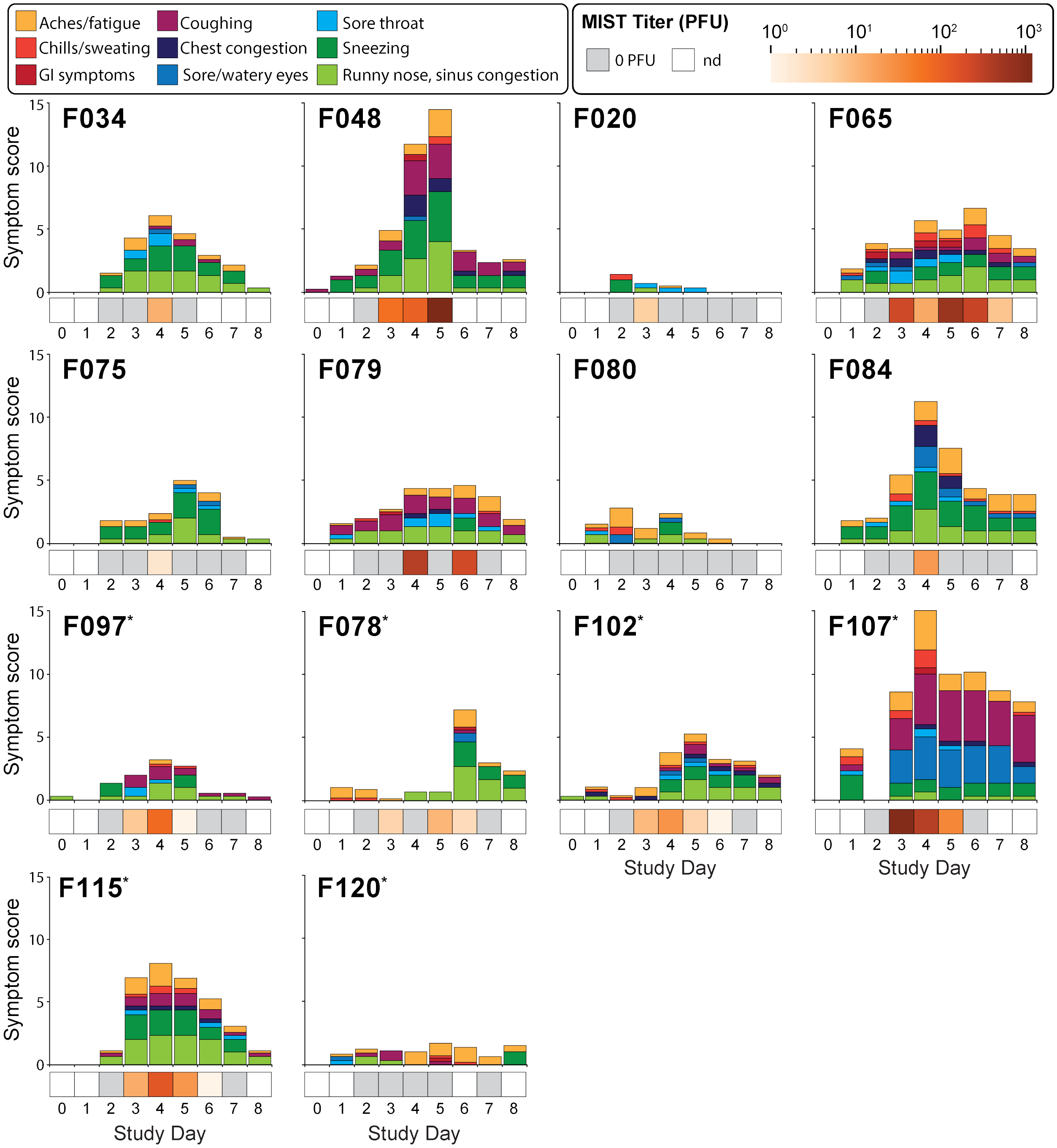


**Supplementary Figure 4. Duration and intensity of symptoms varied across individuals.** Thirty-two symptoms addressed in the FLU-PRO© survey were divided into the nine categories shown in the legend. The score for each category is the mean of the unique symptoms within the category on a scale of 0-4; 0 = "not at all" and 4 = "most severe", that is represented by the height of each stacked, color-coded bar. Symptoms for study day 1 were recorded post-inoculation. Plaque forming units detected in MIST are shown with a heat map underneath each symptom plot. Titers displayed are the sum of plaque forming units detected on all twelve plates in MIST during a given sampling day (inclusive of speaking and coughing and sneezing). Participant identification numbers are shown in the upper left of each facet.

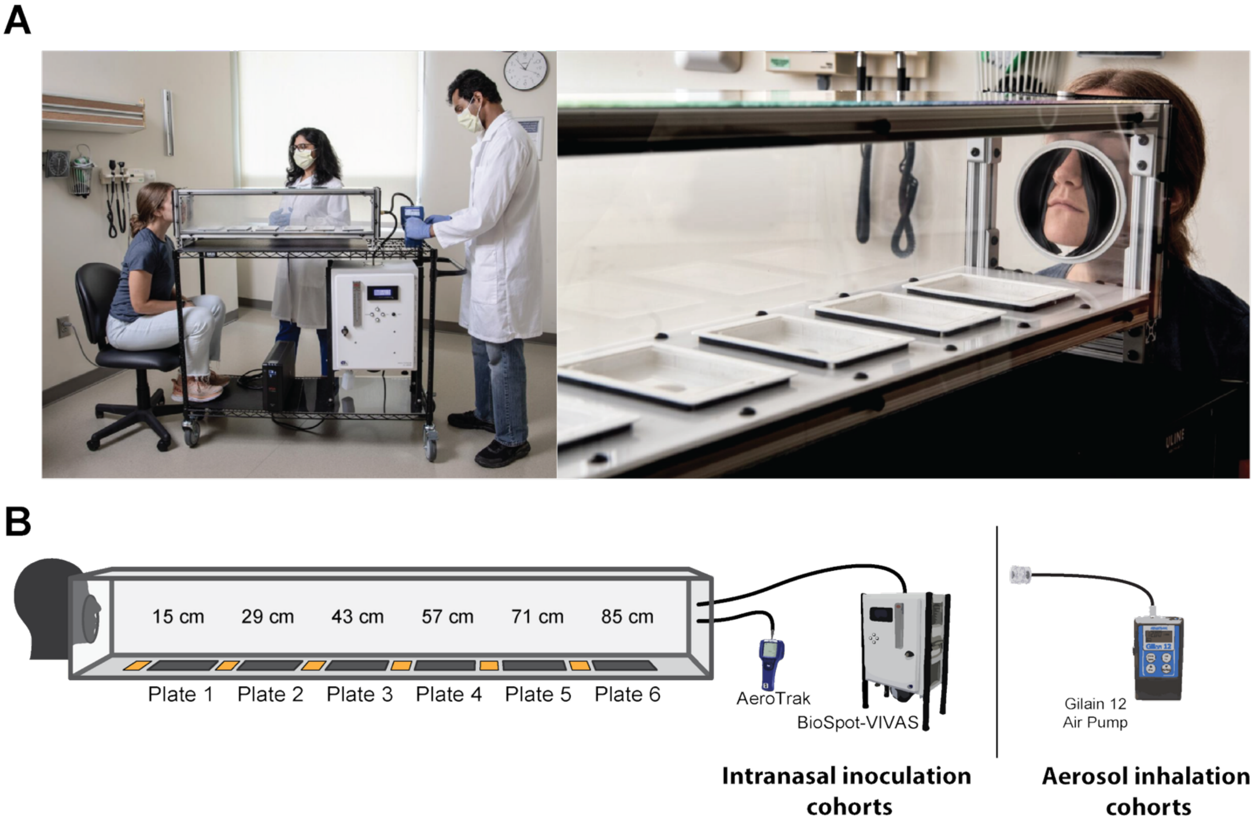


**Supplementary Figure 5. Modular influenza sampling tunnel (MIST).** The MIST is shown schematically, with cell culture plates containing MDCK cells numbered 1-6 and yellow rectangles indicating placement of the water sensitive paper. At the opposite end to the human particle source, air was pulled into AeroTrak and BioSpot VIVAS or the Gilair 12 air pump instruments at rates of 2.8 L/min and 8 L/min, respectively. Staged photos include authors, not participants. Photos are used with the permission of those pictured. Photo credit: Jack Kearse/Emory University.

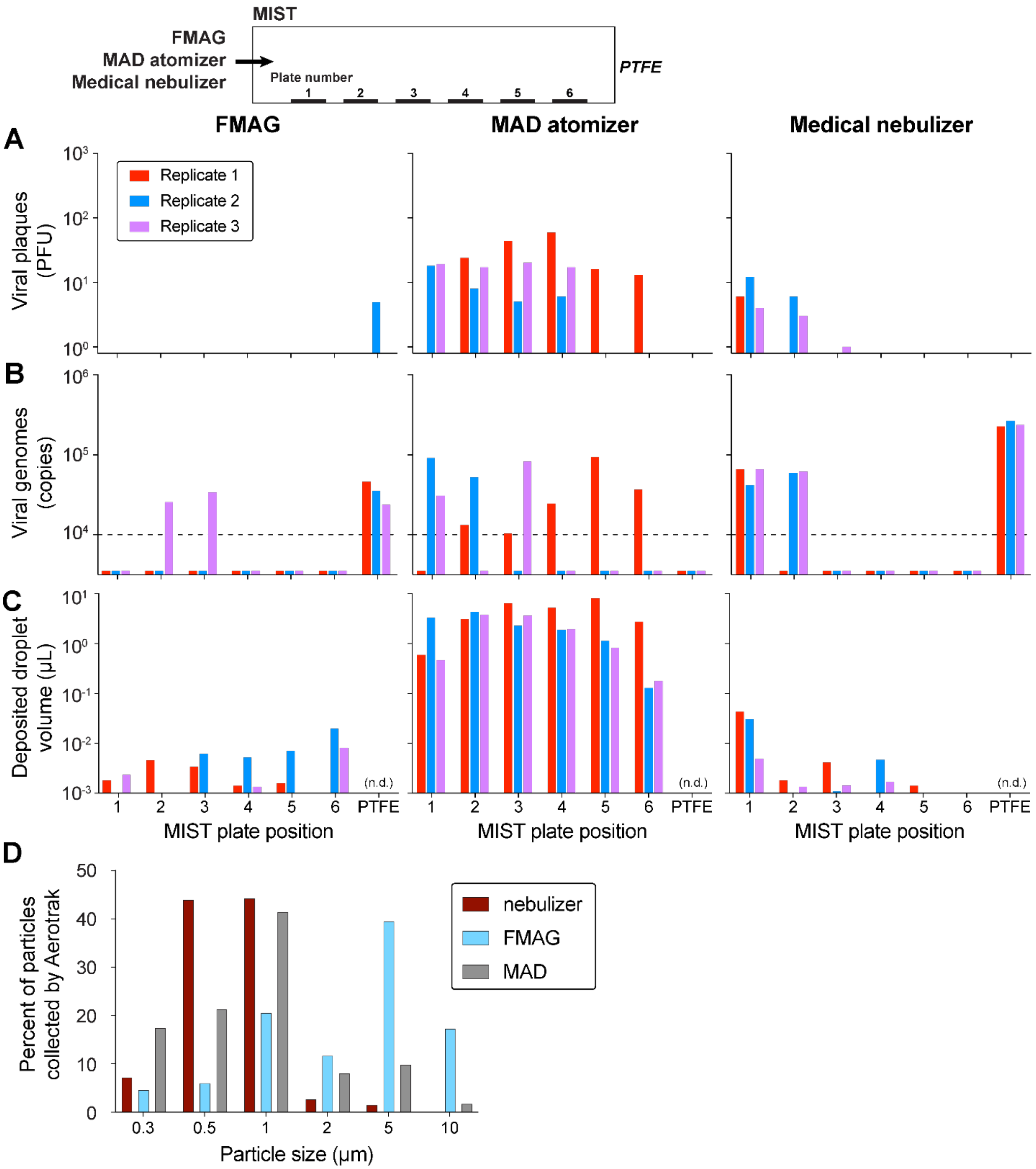


**Supplementary Figure 6. Characterization of particle capture in the MIST.** Synthetic aerosols were generated from 250 µL of a solution containing virus by either 1) medical nebulizer (<2 µm), 2) FMAG (10 µm), or 3) MAD atomizer (30–100 µm). A) Infectious virus detected on each plate in the MIST and terminal PTFE sampler. B) Viral genomes detected on each plate of the MIST and terminal PTFE sampler. Three replicates are shown with different colors.

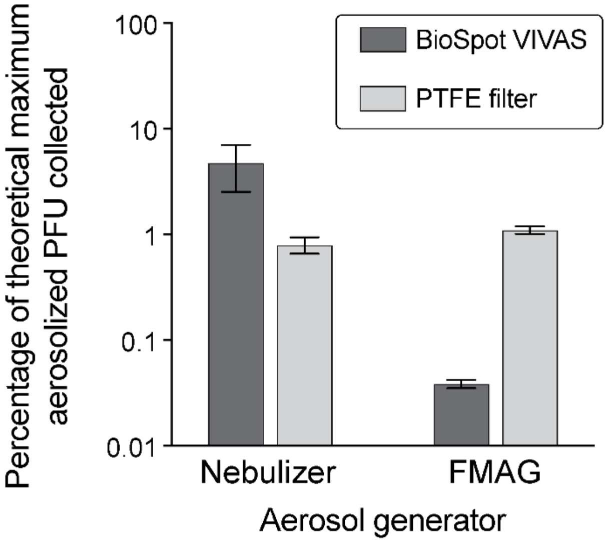


**Supplementary Figure 7. Characterization of particle capture in bioaerosol sampling devices.** Synthetic aerosols containing virus were generated by either 1) medical nebulizer (<2 μm particles) or 2) FMAG (~10 μm particles). The amount of infectious virus captured by the BioSpot Vivas or on PTFE filters was quantified by plaque assay and related to the amount contained in the volume of solution that was aerosolized (the theoretical maximum). Bars show the mean and range of two replicate experiments for the nebulizer and three replicate experiments for the FMAG.

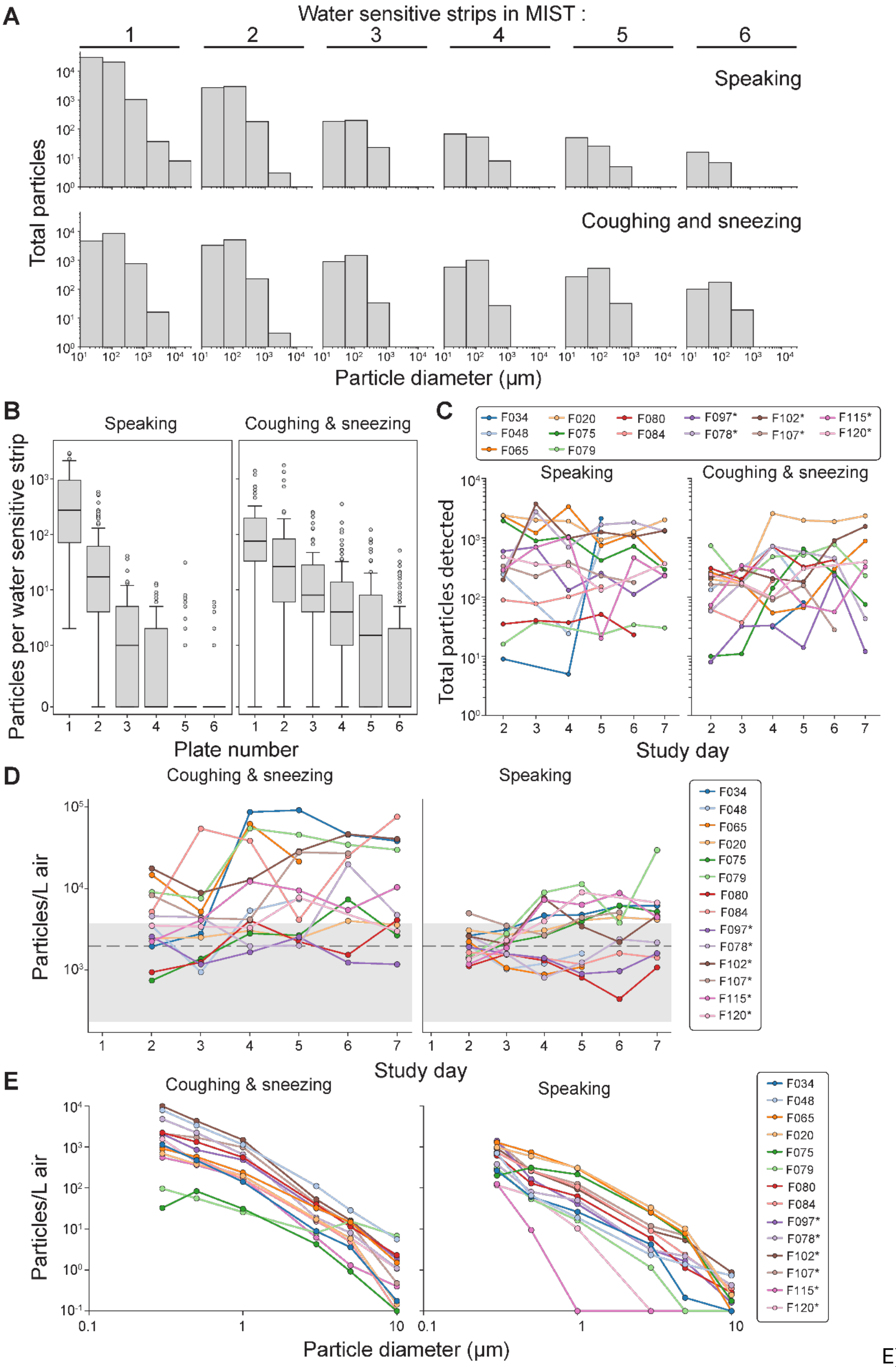
A
Water sensitive strips in MIST :
1 2 3 4 5 6
Speaking
Coughing and sneezing
Total particles
Particle diameter (µm)
B
Speaking
Coughing & sneezing
Particles per water sensitive strip
Plate number
C
F034 F048 F065 F020 F075 F079 F080 F084 F097* F078* F102* F107* F115* F120*
Speaking
Coughing & sneezing
Total particles detected
Study day
D
Coughing & sneezing
Speaking
Particles/L air
Study day
E
Coughing & sneezing
Speaking
Particles/L air
Particle diameter (µm)

E)

**Supplementary Figure 8. Respiratory particles detected in MIST.** A) Size distribution of particles detected on water sensitive strips at six positions in MIST. Data from all participants are combined. Size bins begin at $10^1$ and increment on a $\log_2$ scale. Numbers above the histograms indicate the MIST plate positioned adjacent to the water sensitive strip. B, C) Number of particles detected across all six water sensitive strips in MIST for each participant, plotted over time and according to position in the MIST. Participants were inoculated on Study Day 1. For the boxplots, the central line represents the median, box boundaries indicate the interquartile range, and whiskers extend to data points within 1.5 times the interquartile range. Data points beyond the whiskers are plotted as outliers. D) Particle concentration detected using the AeroTrak sampler at the distal end of MIST for each participant, plotted over time. Participants were inoculated on Study Day 1. Grey shading shows background levels of particles in the air. E) Background subtracted particle counts by particle size collected on Study Day 2 from each participant during speaking or coughing and sneezing using the AeroTrak sampler.

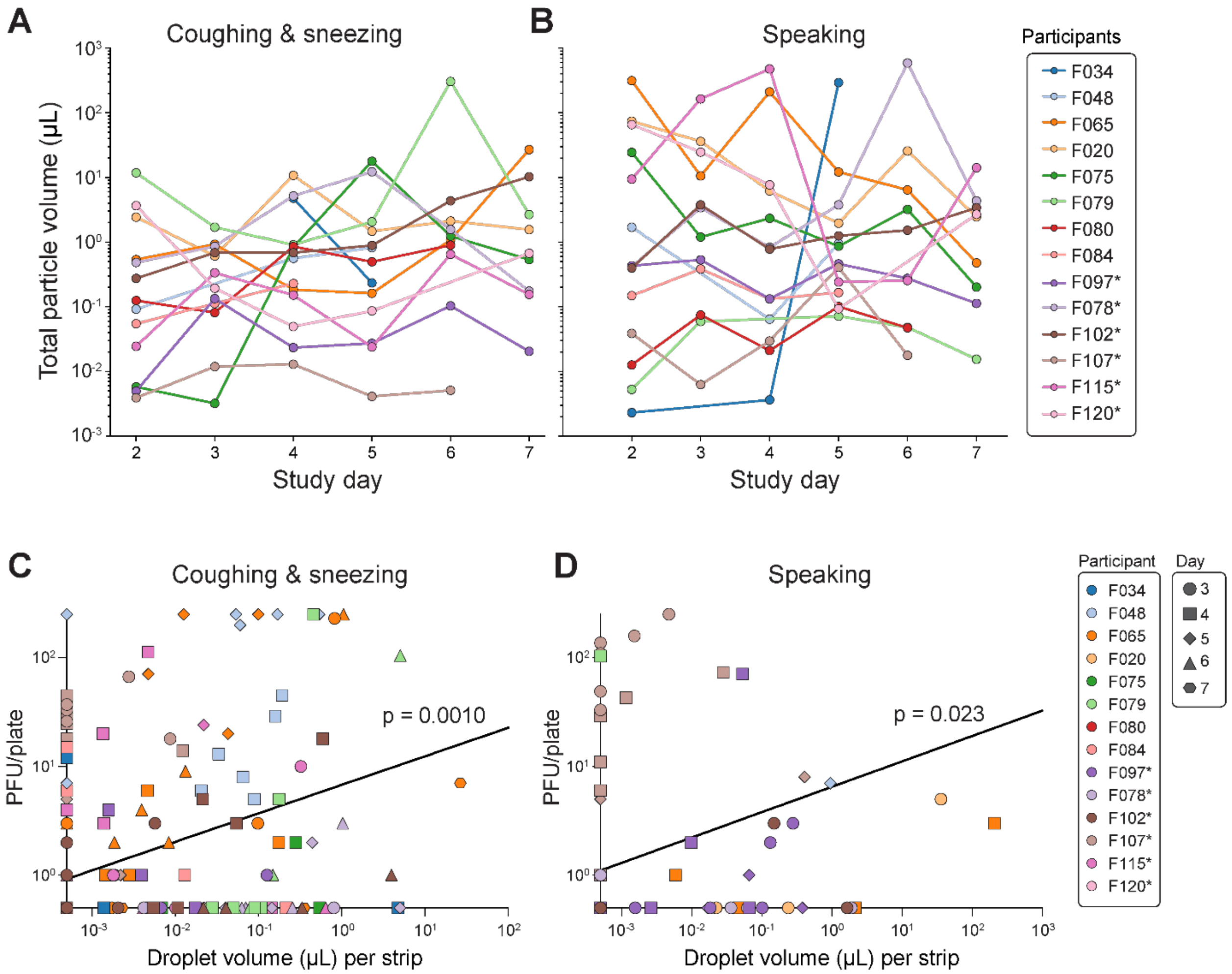


**Supplementary Figure 9. Respired particle volumes are associated with the amount of expelled infectious virus.** The total volume of particles collected on water-sensitive strips within the MIST during (A) coughing and sneezing and (B) speaking varied between individuals and across study days. Linear mixed effects models were used to assess the relationship between PFU collected on each plate and the volume of droplets collected on adjacent strip during (C) coughing and sneezing and (D) speaking using the participant day as a random effect. All plates from days and activities that plaques were detected on at least one plate were included in the analysis. Days and activities where no plaques were detected were not included. Data were log transformed prior to statistical analysis. Each participant is color coded with days shown with different shapes. The black lines are linear regressions from the model. The p-values indicate the significance of the association between the two fixed-effects variables. Data points with no measurable values were set to half of the limits of detection for each measurement (0.5 PFU per MIST plate and $5x10^{-4}$ for droplet volume) for the purpose of this analysis. Plates with too many PFU to count were assigned values of 250 PFU. F048 day 3 coughing/sneezing, and F102 day 5 speaking and coughing/sneezing did not have droplet volume data collected at the same time that plaques were detected so they were not included in the analysis.

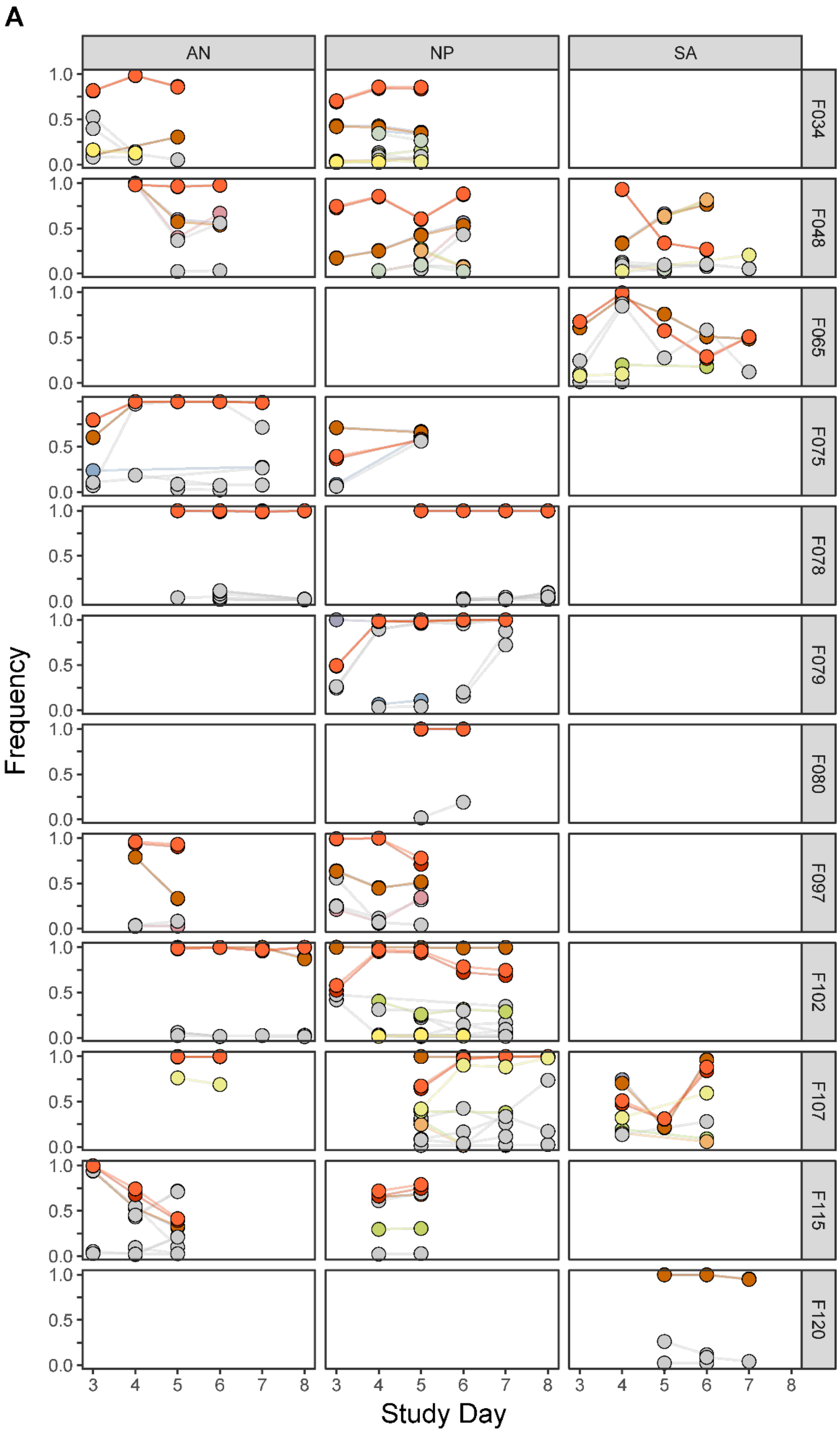
A
AN
NP
SA
F034
F048
F065
F075
F078
F079
F080
F097
F102
F107
F115
F120
1.0
0.5
0.0
3
4
5
6
7
8
Frequency
Study Day

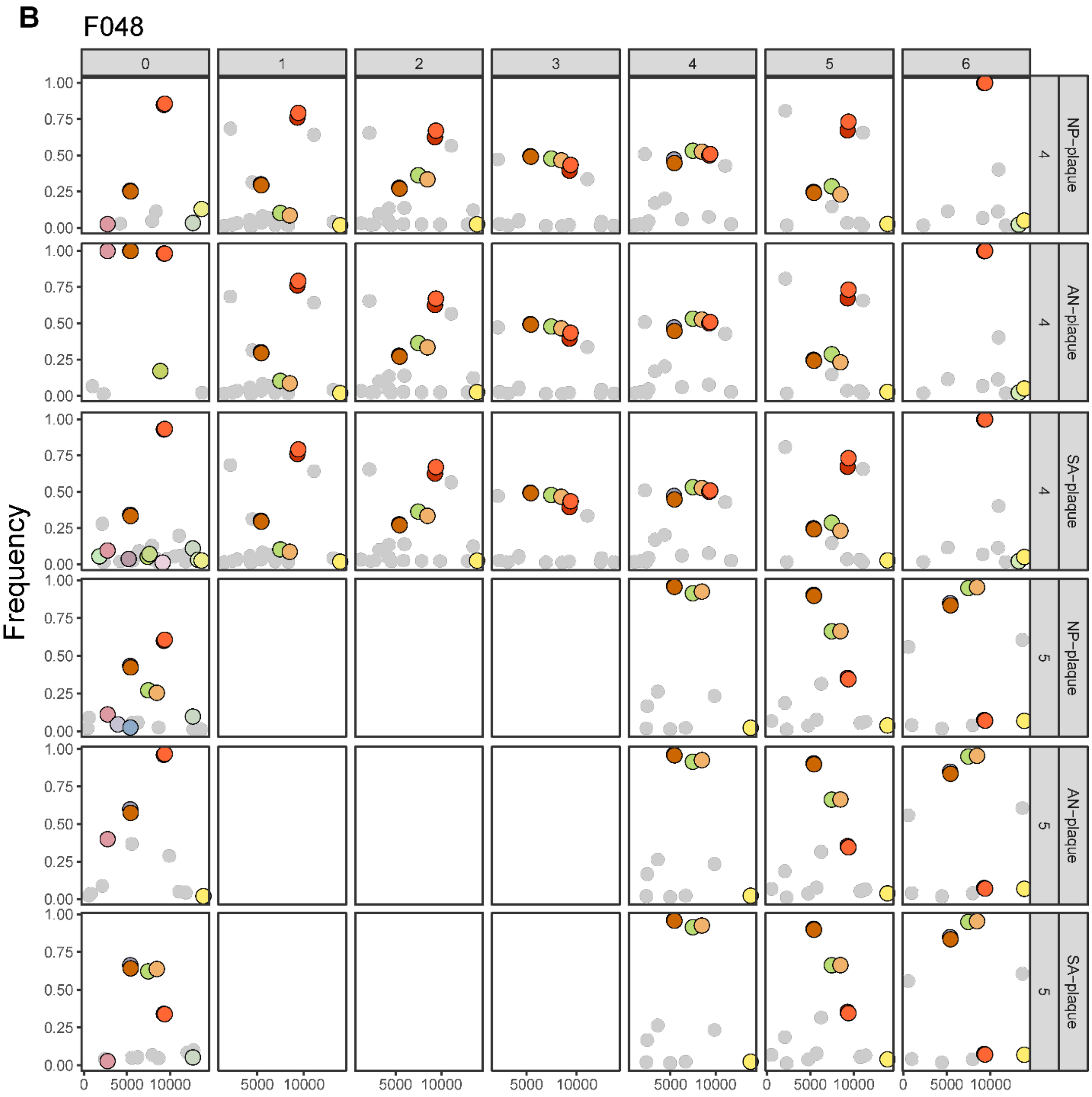

B
F048
0
1
2
3
4
5
6
NP–plaque
AN–plaque
SA–plaque
4
5
Frequency
1.00
0.75
0.50
0.25
0.00
0
5000
10000

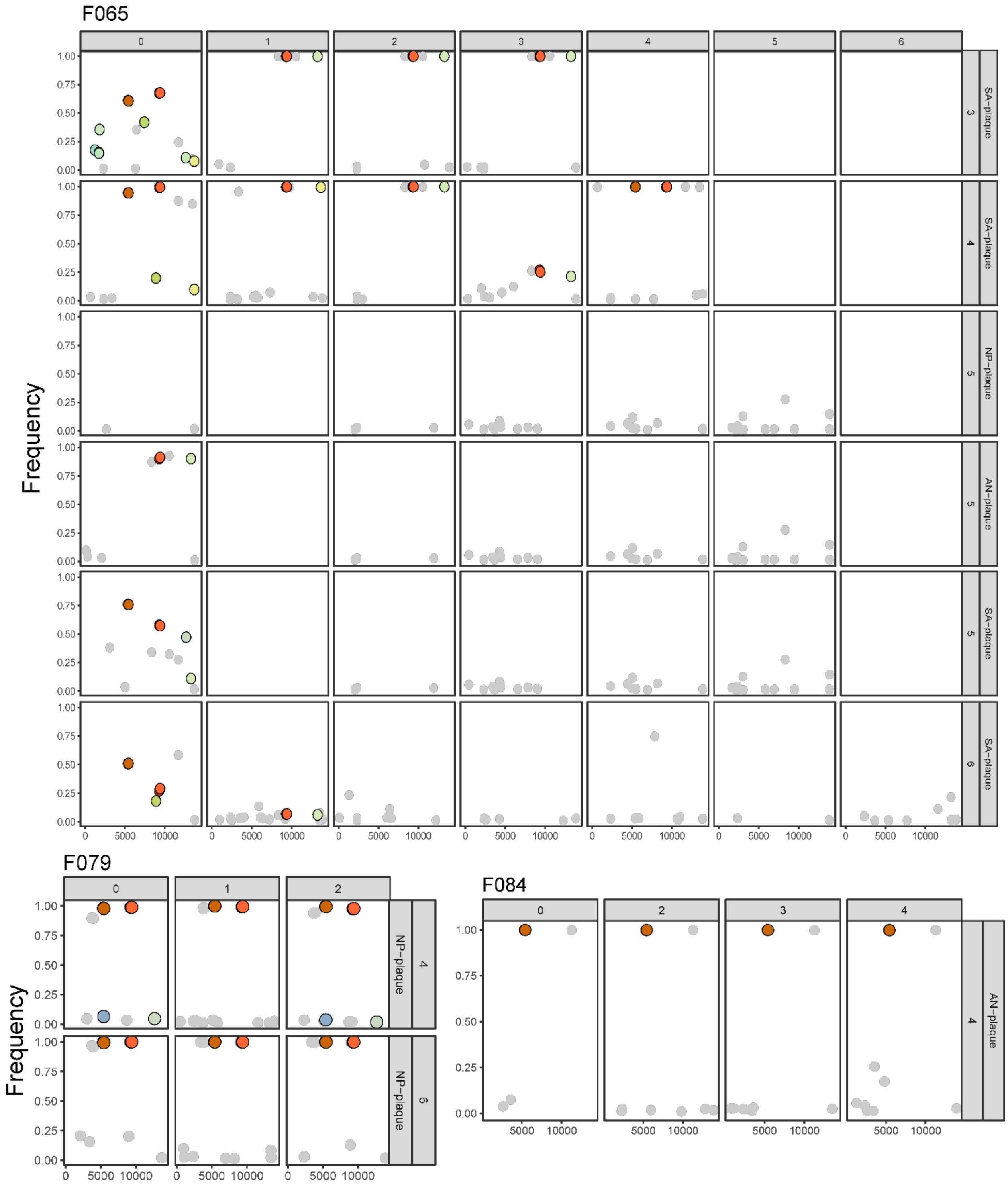

F065
0
1
2
3
4
5
6
SA−plaque
3
SA−plaque
4
NP−plaque
5
AN−plaque
5
SA−plaque
5
SA−plaque
6
Frequency
1.00
0.75
0.50
0.25
0.00
0
5000
10000
F079
NP−plaque
4
NP−plaque
6
F084
AN−plaque
4

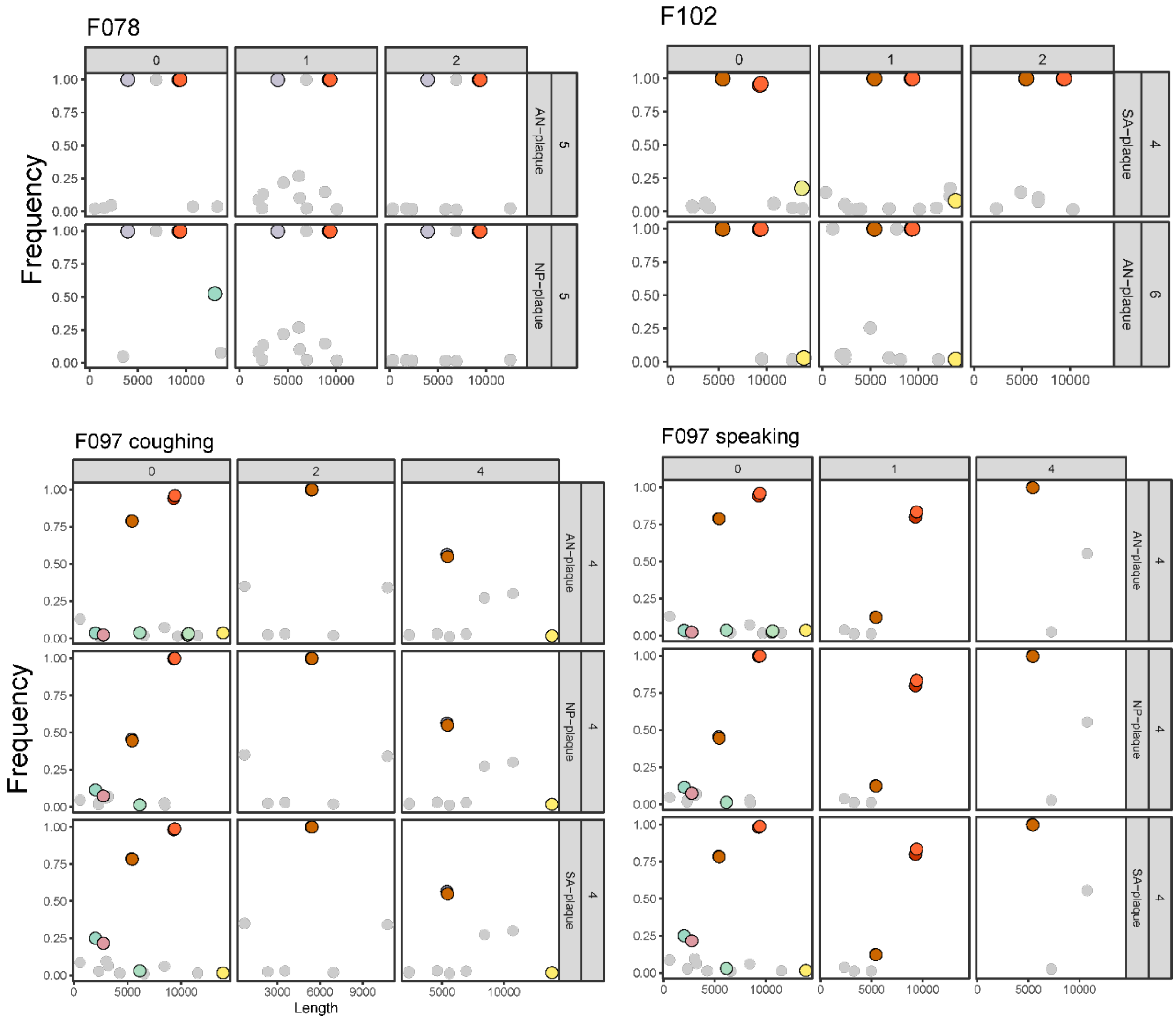
F078
0
1
2
AN−plaque
5
NP−plaque
5
Frequency
1.00
0.75
0.50
0.25
0.00
0
5000
10000
F102
0
1
2
SA−plaque
4
AN−plaque
6
F097 coughing
0
2
4
AN−plaque
4
NP−plaque
4
SA−plaque
4
3000
6000
9000
Length
F097 speaking
0
1
4

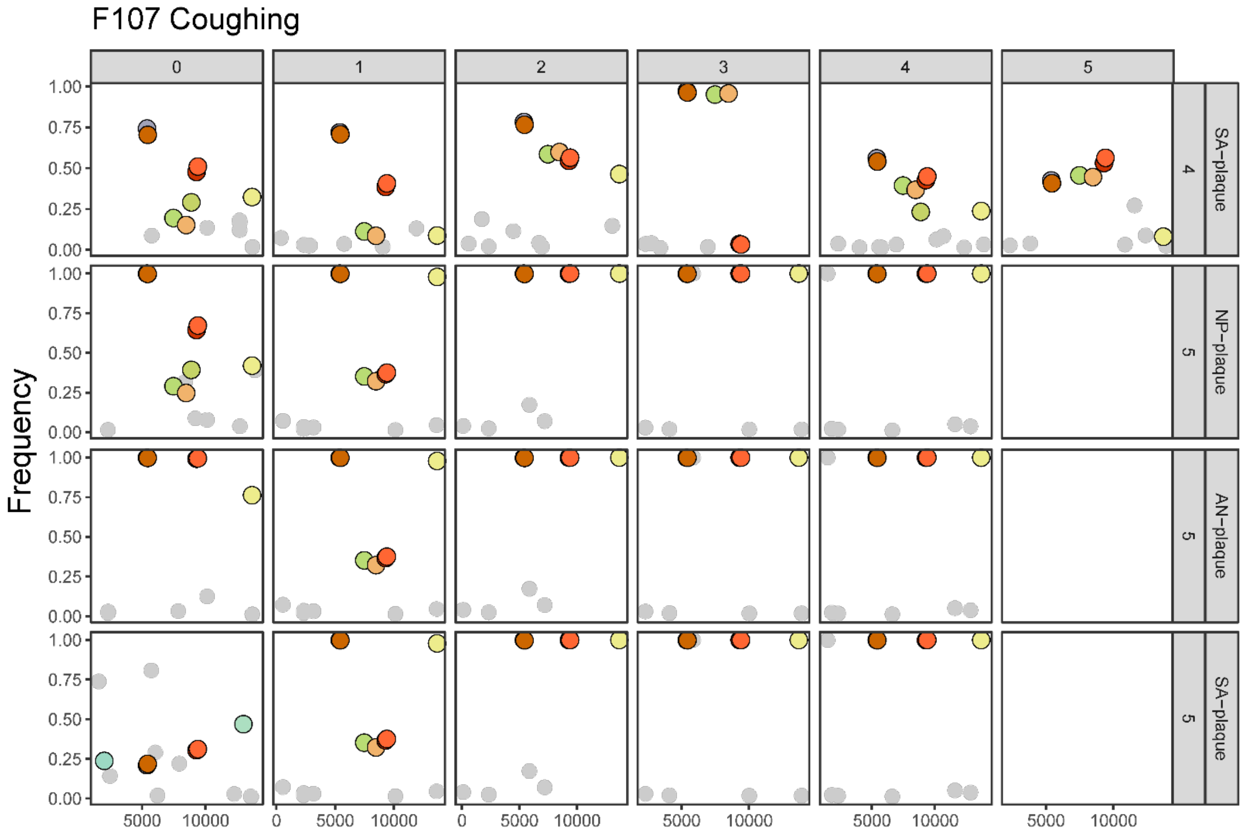
F107 Coughing
Frequency
0
1
2
3
4
5
SA–plaque
4
NP–plaque
5
AN–plaque
5
SA–plaque
5
1.00
0.75
0.50
0.25
0.00
5000
10000
0

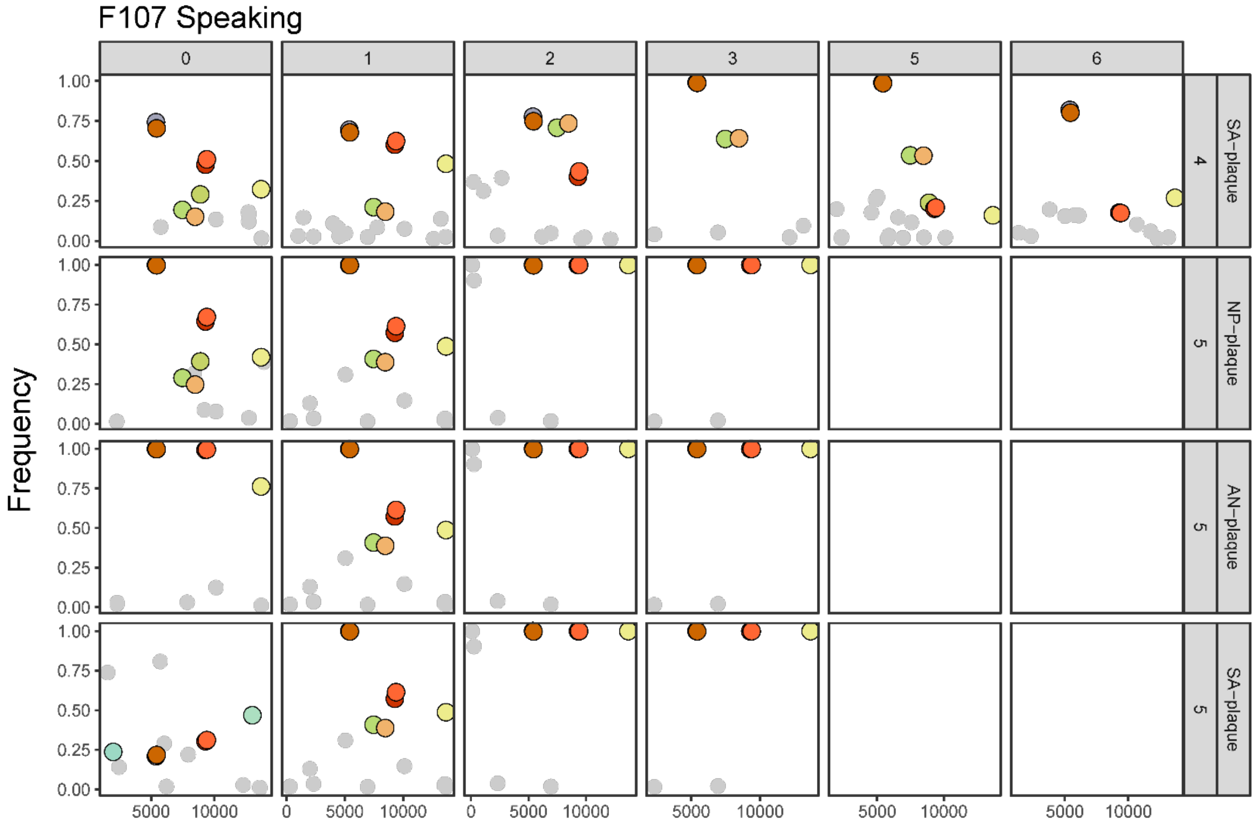
F107 Speaking
Frequency
0
1
2
3
5
6
SA–plaque
4
NP–plaque
5
AN–plaque
5
SA–plaque
5
1.00
0.75
0.50
0.25
0.00
5000
10000
0

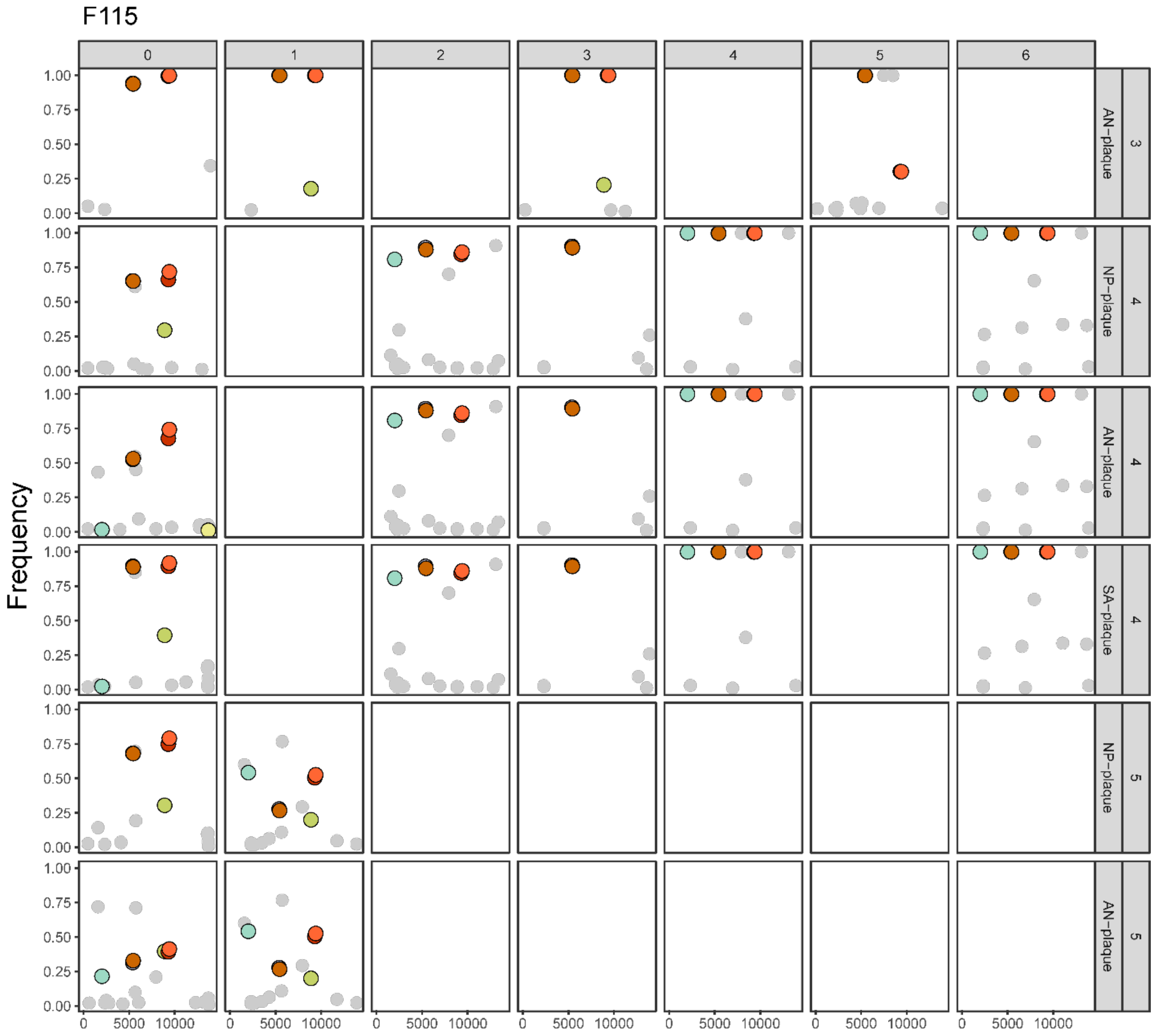


**Supplementary Figure 10. Carry-over of variants from the inoculum to participants.** A) Variant frequencies over time observed in participants. Only variants detected in the inoculum are shown, with a specific color assigned to each unique variant. Sample type is shown at the top. NP=nasopharyngeal swab; AN=anterior nasal swab; and SA=saliva. Participant ID is shown at the right. B) Each panel represents samples from a unique individual over several study days. Within-host variants are reported in the first column of every panel (labeled 0), with sample type and study day shown at the right. Variants detected in MIST are reported in grids labeled 1-6, where 1 represents the culture plate positioned closest to the source. Colored data points represent variants found in the inoculum, with a specific color assigned to each unique variant. Grey circles represent variants found exclusively in the MIST. The y-axis variant frequency. The x-axis shows nucleotide position in a concatenated version of the influenza A virus genome.

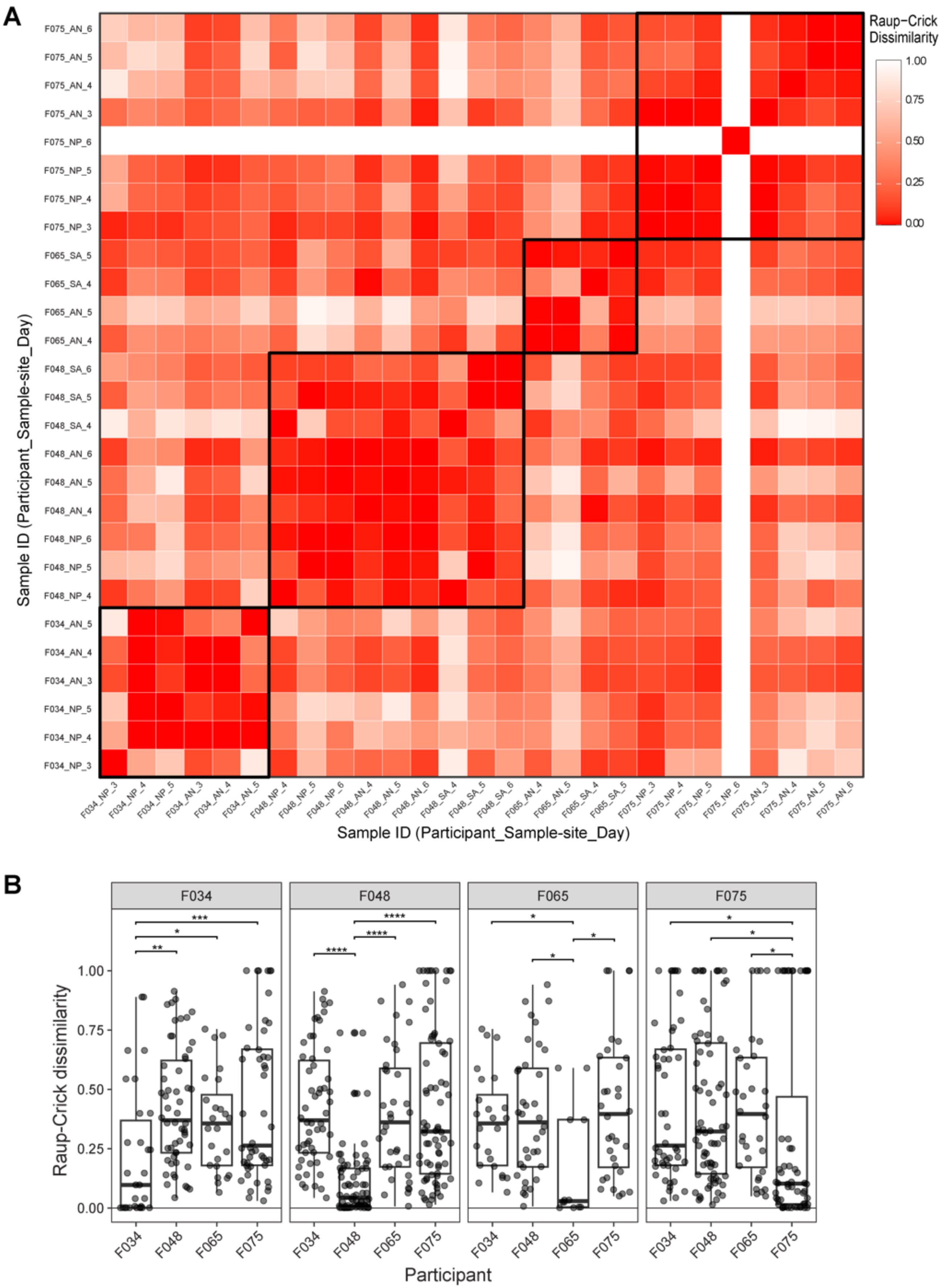
A
B
Raup−Crick Dissimilarity
1.00
0.75
0.50
0.25
0.00
Sample ID (Participant_Sample-site_Day)
F075_AN_6
F075_AN_5
F075_AN_4
F075_AN_3
F075_NP_6
F075_NP_5
F075_NP_4
F075_NP_3
F065_SA_5
F065_SA_4
F065_AN_5
F065_AN_4
F048_SA_6
F048_SA_5
F048_SA_4
F048_AN_6
F048_AN_5
F048_AN_4
F048_NP_6
F048_NP_5
F048_NP_4
F034_AN_5
F034_AN_4
F034_AN_3
F034_NP_5
F034_NP_4
F034_NP_3
Sample ID (Participant_Sample-site_Day)
F034
F048
F065
F075
Raup-Crick dissimilarity
1.00
0.75
0.50
0.25
0.00
Participant

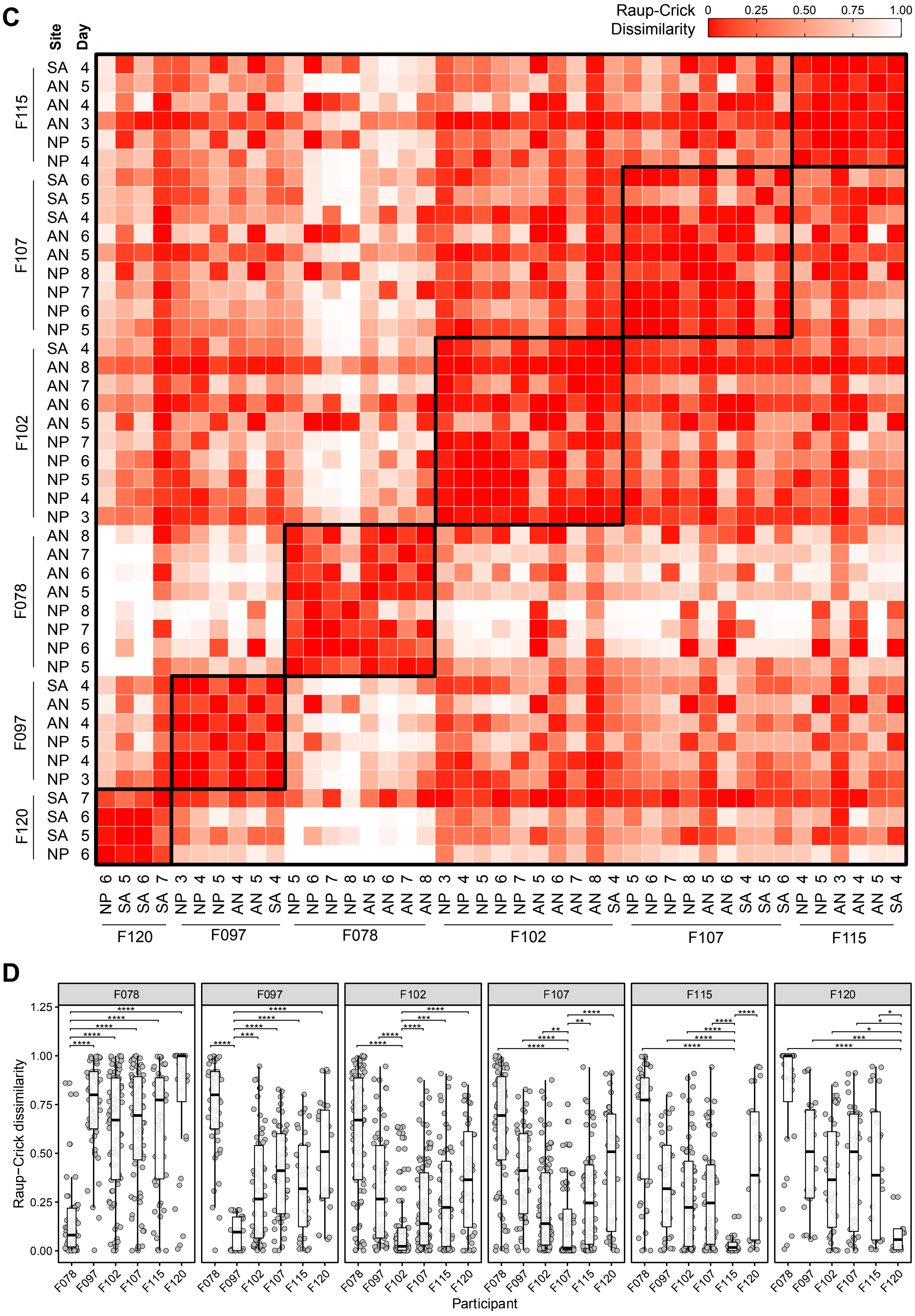
C
Raup-Crick
Dissimilarity
0
0.25
0.50
0.75
1.00
Site
Day
F115
F107
F102
F078
F097
F120
SA 4
AN 5
AN 4
AN 3
NP 5
NP 4
SA 6
SA 5
SA 4
AN 6
AN 5
NP 8
NP 7
NP 6
NP 5
SA 4
AN 8
AN 7
AN 6
AN 5
NP 7
NP 6
NP 5
NP 4
NP 3
AN 8
AN 7
AN 6
AN 5
NP 8
NP 7
NP 6
NP 5
SA 4
AN 5
AN 4
NP 5
NP 4
NP 3
SA 7
SA 6
SA 5
NP 6
D
F078
F097
F102
F107
F115
F120
Raup−Crick dissimilarity
0.00
0.25
0.50
0.75
1.00
1.25
Participant

**Supplementary Figure 11. Viral populations sampled across time and locations within one participant show lower dissimilarity than those sampled from unrelated participants.** A and C) Raup-Crick dissimilarity index comparison between participants, samples and study day. Black boxes delimit comparisons made within a given participant. B and D) Raup-Crick dissimilarity index pairwise comparison between participants. Shapiron-Wilks normality test determined that dissimilarity followed a nonnormal distribution (W= 0.89. p<2.2e-16). Therefore, the Wilcoxon Rank-Sum test was performed to determine statistical significance. Statistical results are shown only for those that were significant. F034-F048: p=9.42e-04; F034-F065: p=2.60e-02; F034-F075: p=3.00e-03; F048-F034: p=1.15e-13; F048-F065: p=1.65e-09; F048-F075: p=1.82e-12; F065-F034: p=3.70e-02; F065-F048: p=1.80e-02; F065-F075: p=2.30e-02; F075-F034: p=6.00e-03; F075-F048: p=6.00e-03; F075-F065: p=2.30e-02

### Intranasal inoculated

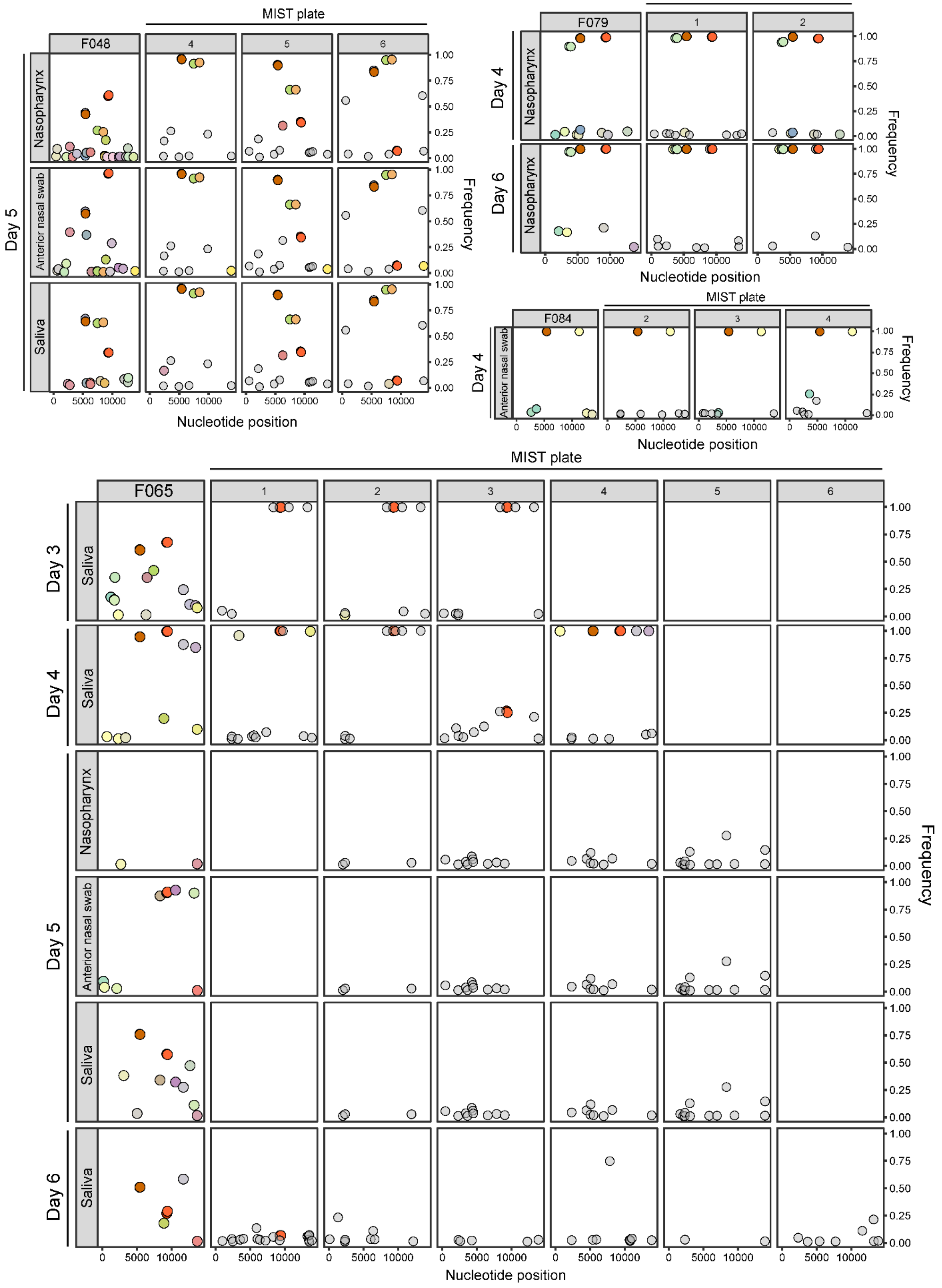

### Aerosol inoculated

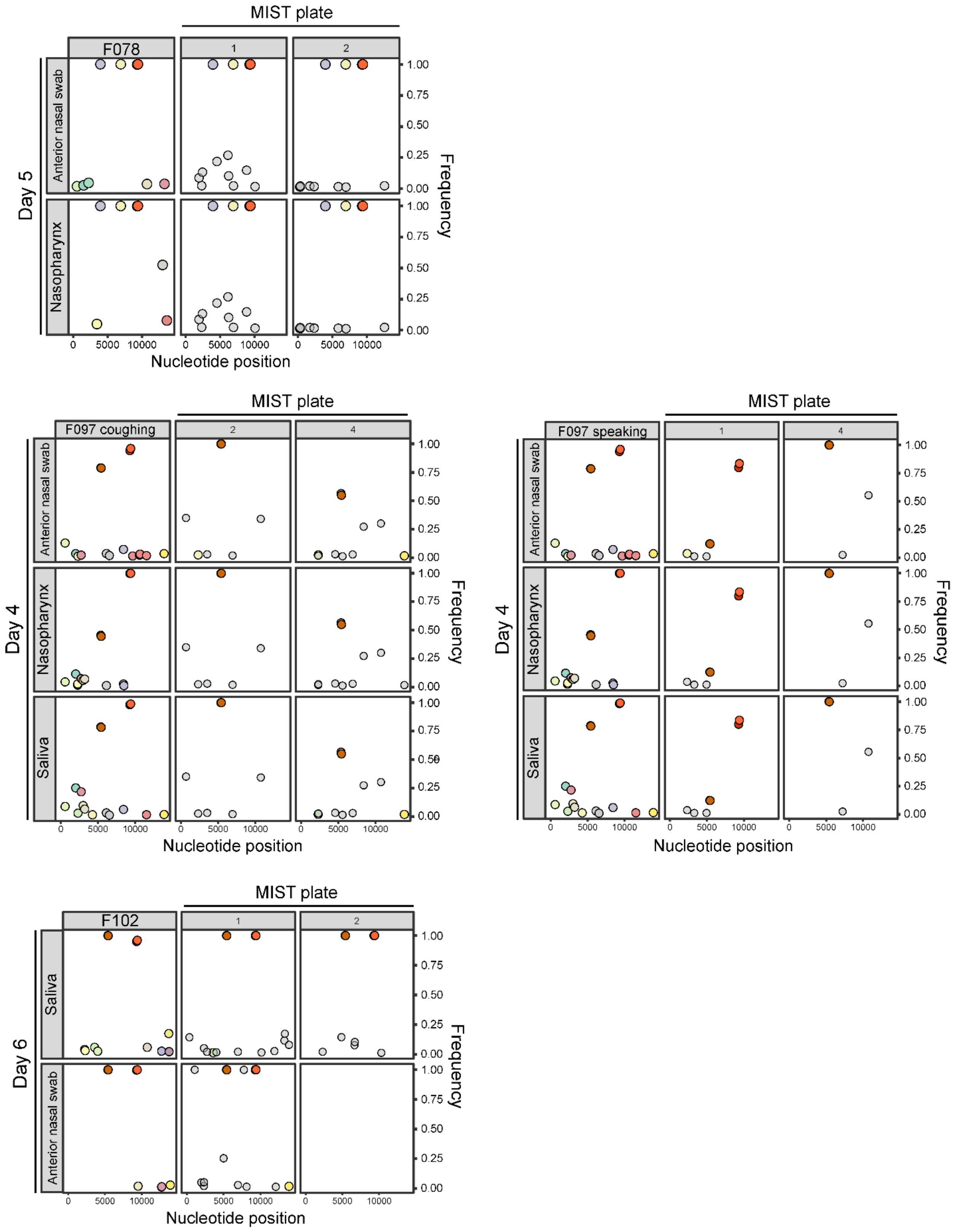

### Aerosol inoculated

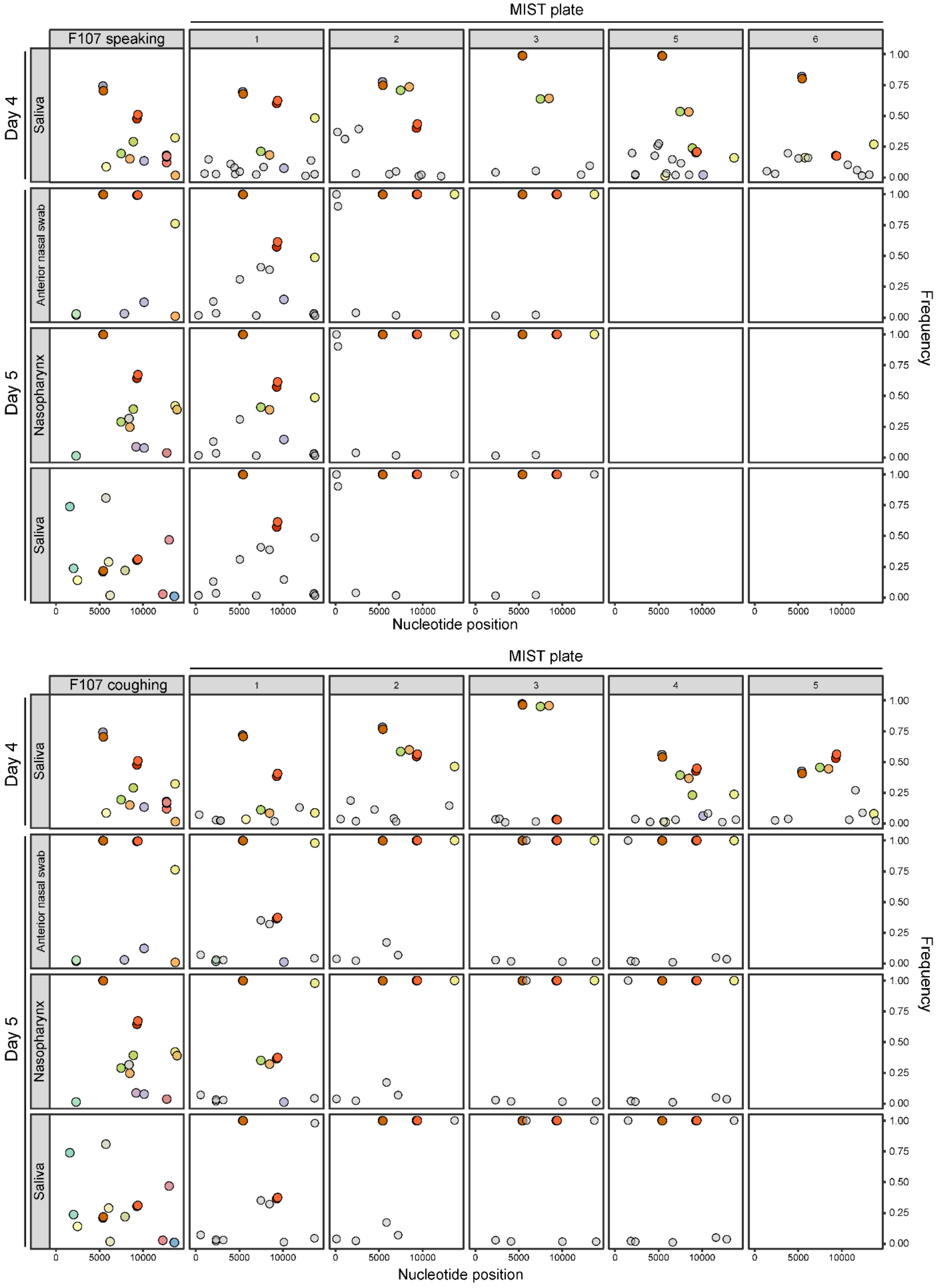

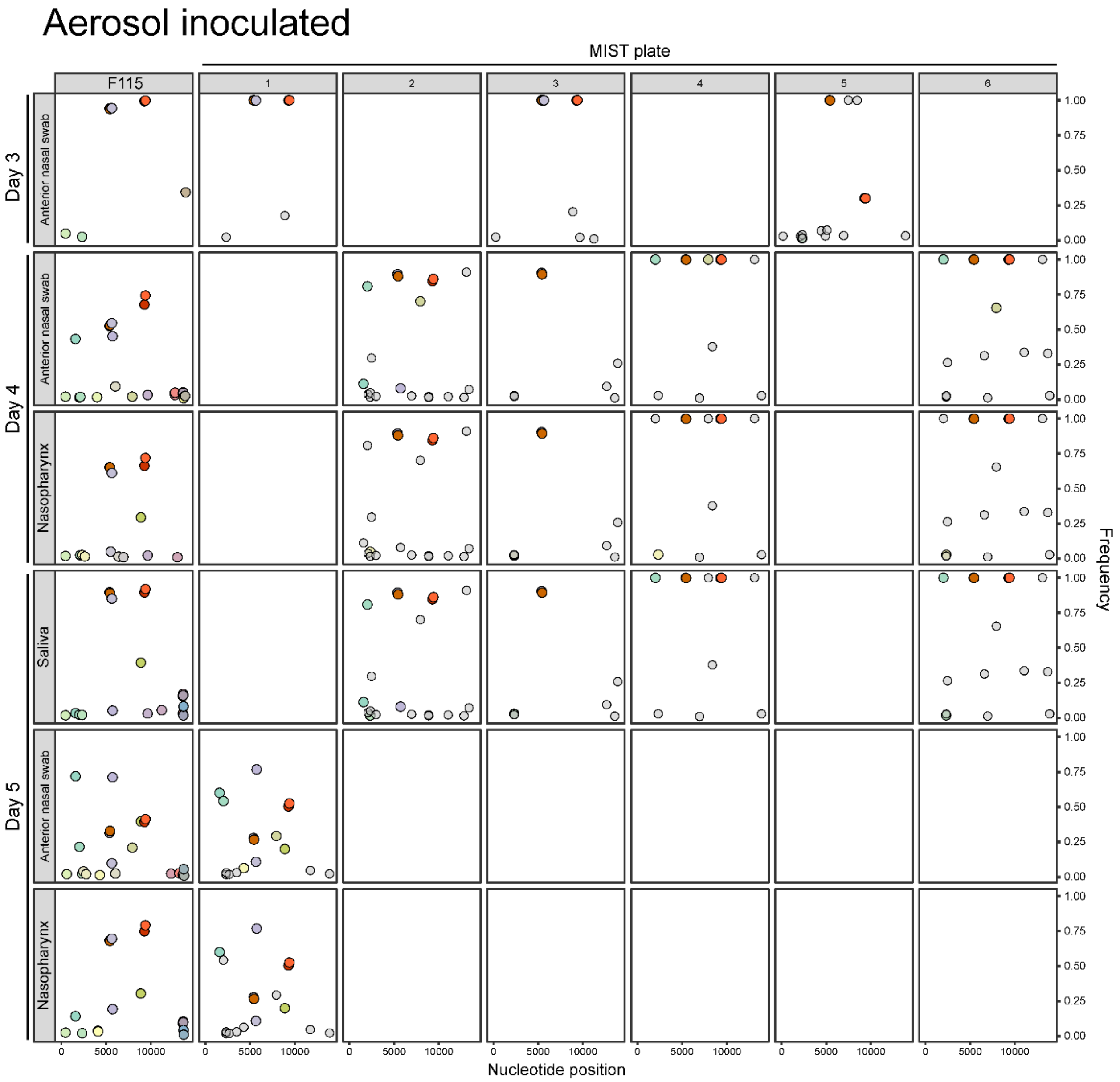


**Supplementary Figure 12. Genetic composition of viral populations in infectious respiratory particles produced during coughing and sneezing.** Each panel represents samples from a unique individual over several study days. Within-host variants in the nasopharynx (NP swabs), anterior nasal swabs, and saliva are reported in the first column of every panel. Variants detected in MIST are reported in grids labeled 1-6, where 1 represents the culture plate positioned closest to the source. Colored data points represent variants found within the host, with a specific color assigned to each unique variant. Grey circles represent variants found exclusively in MIST. The y-axis on the right shows variant frequency. The x-axis shows nucleotide position in a concatenated version of the influenza A virus genome.

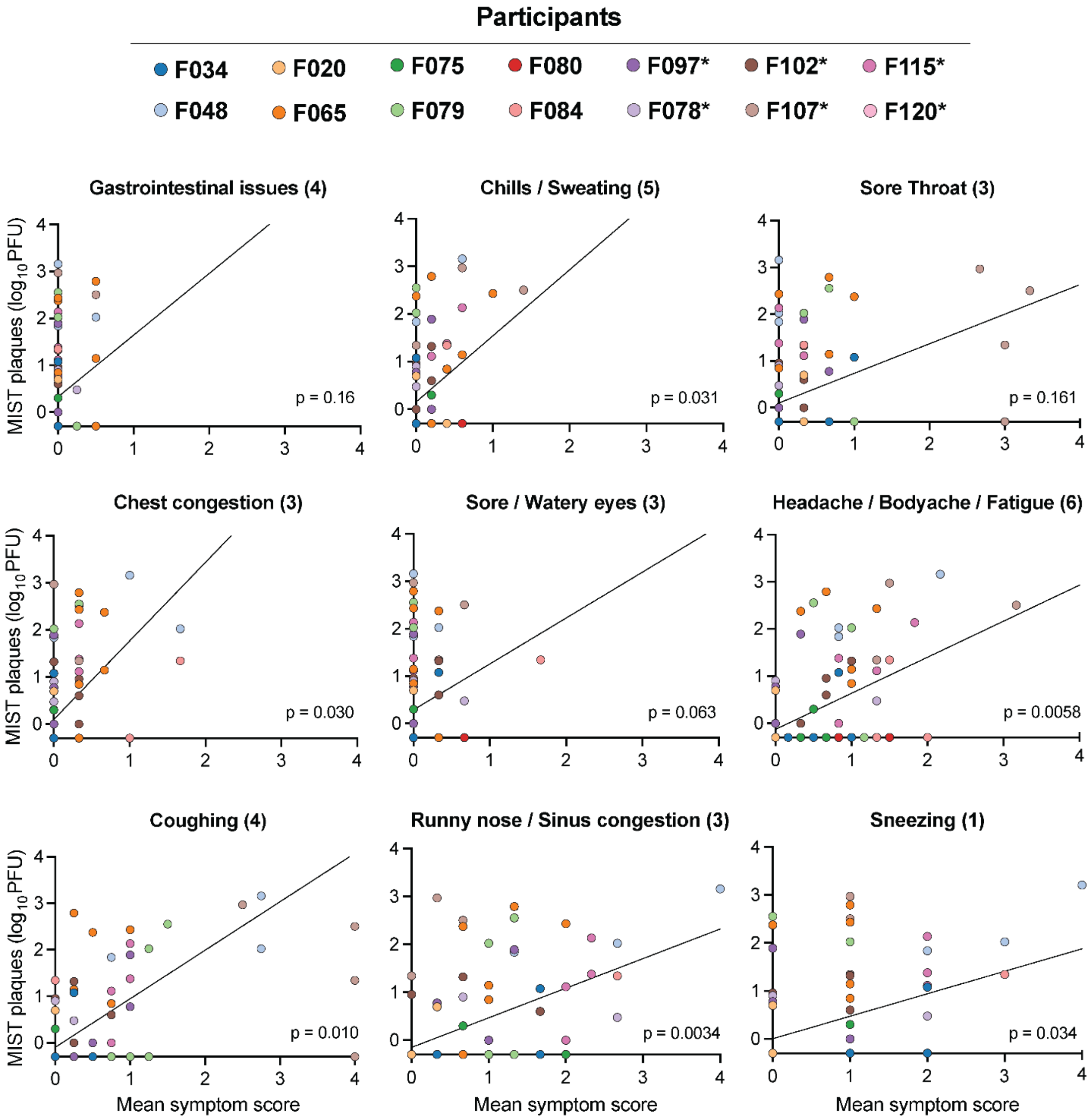


**Supplementary Figure 13. Relationships between symptoms and expulsion of infectious virus.** Daily mean symptom scores for each symptom category are plotted against the base-10 logarithm of the total number of plaques counted on all plates from speaking and coughing and sneezing into the MIST on the corresponding study day. The number of symptoms in each category is indicated in parentheses, with each symptom rated on a severity scale that ranges from 0-4; ( "not at all" – "most severe" ). Linear mixed effects models of the relationships between infectious titers in the MIST and mean symptom scores are shown. Regressions from each model are shown using black lines. p-values indicate the significance of the association between the two variables. Each participant is indicated using a different color. Undetectable MIST plaques were set to 0.5 (half of the limit of detection). n = 78 study days. Missing symptom responses were not included in calculations of mean symptom scores.

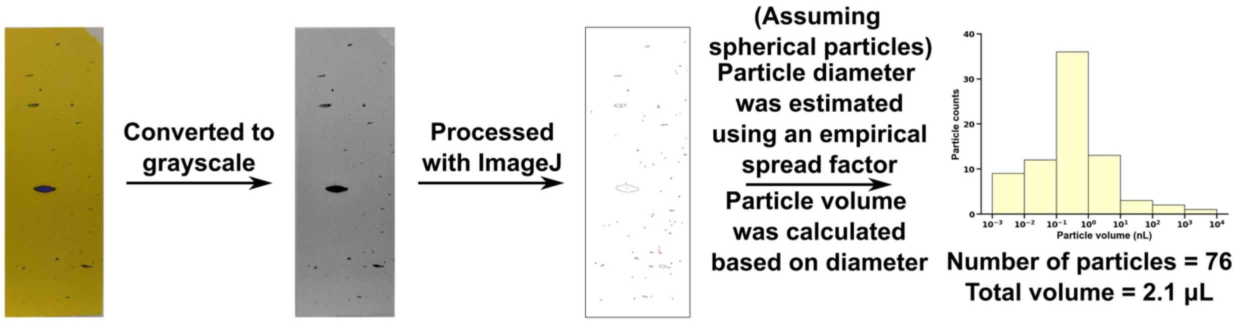


**Supplementary Figure 14. Analysis of water-sensitive strips.** The image of the water-sensitive strip is first converted to grayscale. The grayscale image is then processed using ImageJ software to identify the size and number of deposited particles. An empirical correction factor is applied to the identified particle sizes to account for the spreading of particles on the water-sensitive strips. The corrected diameters were used to determine the total volume of particles on each strip.
